# Clinical effectiveness of computer-based psychoeducational self-help platforms for eating disorders (with or without an associated app): A systematic review

**DOI:** 10.1101/2024.11.08.24316381

**Authors:** Alessandra Diana Gentile, Yosua Yan Kritian, Erica Cini

**Affiliations:** Department of Child & Adolescent Psychiatry, Institute of Psychiatry, Psychology & Neuroscience, King’s College London, London, United Kingdom; Division of Medicine, University College London, London, United Kingdom; East London NHS Foundation Trust, London, United Kingdom

**Keywords:** self-help, eating disorders, disordered eating, prevention, digital health, psychoeducation, computer-based

## Abstract

**Background:** Following the COVID-19 pandemic, computer-based self-help platforms for eating disorders (EDs) became increasingly prevalent as a tool to effectively prevent and treat ED symptoms and related behaviours. This systematic review explored the clinical effectiveness of computer-based self-help platforms for EDs.

**Methods:** From inception to the 31^st^ of May 2024, a systematic search of Ovid MEDLINE, Embase, Global Health, and APA PsycInfo was conducted. This systematic review followed the Preferred Reporting Items for Systematic Reviews and Meta-Analyses (PRISMA) guidelines. Outcome quality assessments were conducted according to the Grading of Recommendations Assessment, Development and Evaluation (GRADE).

**Results:** 14 RCTs, with a total of 4195 participants, were included. 4 studies explored the effectiveness as primary prevention, 7 as secondary prevention, and 3 as tertiary intervention. The gathered literature demonstrated computer-based self-help platforms as clinically effective in reducing ED core symptoms and related behaviours, with psychoeducation, cognitive behavioural, and dissonance-based approaches being the most prevalent approaches.

**Conclusions:** Computer-based self-help platforms are effective in the short-term reduction of ED symptoms and associated behaviours and should be implemented in the early stages of a tiered healthcare system for ED treatments.

**Trial Registration:** Prospero (CRD42024520866).

## Introduction

Eating disorders (ED) are a group of psychiatric disorders characterised by irregular eating behaviours, body image concerns, and, in some types, fear of gaining weight [1]. Literature shows that 5.5% to 17.9% of females and 0.6% to 2.4% of males are diagnosed with EDs based on Diagnosis and Statistical Manual of Mental Disorders (DSM-5) criteria [2].

Moreover, some minority groups are at particular risk of developing EDs. For example, gender and sexual minorities are demonstrated to be at greater risk for developing EDs, with anorexia nervosa and bulimia being the most prevalent [2–5]. Notably, compared to any other psychiatric condition, anorexia nervosa has the highest suicidality and mortality rates, and lowest quality of life levels, highlighting the importance for urgency of care [6, 7].

Weissman and Roselli (2017) stated that merely 25% of individuals with ED symptoms or developed EDs access care, which is explained by the lack of accessible treatment and individual treatment preferences [8]. EDs have been demonstrated to negatively impact the psychological, cognitive, physical, and social development of individuals, which evidences the need for accessible resources which target early identification through psychoeducation, such as self-help platforms, which support individuals through early intervention and resource signposting [9].

Psychoeducation improves recognition by identifying the symptoms and warns patients about the negative impact of EDs on their physical and mental health, which in turn increases awareness and overall demystifies the disorder [9, 10]. This highlights that psychoeducation is an effective tool for the prevention of EDs and promoting well-being among the public [11, 12]. Self-help platforms support people’s understanding of their symptoms, and some platforms help with the development of coping skills, through for example, cognitive behavioural therapy (CBT), dissonance-based intervention (DBI), motivational-enhancement therapy (MET)-in form techniques, as well as directing people to seek early help [13–15].

Self-help platforms mitigate barriers such as large geographical distances from healthcare clinics, treatment-seeking stigma, and time constraints, whilst also reducing ongoing staff-related treatment costs [16, 17]. Further, COVID-19 put ED services under amplified pressure due to the increase in referrals and acuity [18]. This places self-help platforms in good stead to provide support to the general public and to those who require early intervention. It also has a role in creating a ‘waiting well’ environment, which refers to a proactive approach in which individuals and their families engage in purposeful activities, such as self-guided psychoeducation to manage and mitigate the impacts of EDs and disordered eating behaviours (DEBs)[19, 20, 21]. Overall, this ensures progress and better symptom management, reduced anxiety, and planning ahead prior to engaging with formal interventions [21].

In line with the National Institute for Health and Care Excellence (NICE) guidelines, ED interventions follow the stepped care model [22]. The stepped care model suggests that individuals should first receive low-intensity interventions such as psychoeducation and progressively receive more intensive interventions if necessary [23]. This aligns with the THRIVE framework, which highlights a needs-led approach to delivering person-centred integrated interventions [24]. According to the THRIVE framework, significance is placed on the prevention of mental health issues and the promotion of wellbeing in the general public. Computer-based self-help platforms ± associated apps allow for accessible and scalable support, taking into consideration the unlimited demand and the limited resources [25–27]. Various digital modalities have been tried, such as compact disc read-only memory (CD-ROM), videos, and text messages, but no strong evidence emerged [28, 29]. Self-help platforms which utilise internet cognitive behavioural therapy (ICBT or CBI-I) and internet dissonance-based intervention (IDBI) have been demonstrated to be feasible and effective alternatives [30, 31]. There is evidence that computer-based self-help platforms ± associated apps effectively reduce ED symptoms and ED-related behaviours [32, 33].

Self-help platforms can be utilised as part of therapy using a guided approach. A guided approach involves the support of a clinician or peers while the user navigates the platform and learns accurate information. Systematic reviews which compared guided to non-guided self-help platform effectiveness in reducing ED symptomology found that guided interventions are significantly more effective compared to nonguided self-help interventions [11, 34]. Additionally, guided self-help platforms were shown to significantly increase intervention adherence and participant satisfaction [35]. On the contrary, Aardom (2017) showed that an unguided computer-based intervention is more suitable for patients with EDs who exhibit mild to moderate bulimic symptoms, but less effective for those with severe symptoms [32]. This can be explained by the mild and moderate symptomologies requiring less intensive support; therefore, allowing individuals to manage their symptoms more effectively. Contrastingly, severe symptomologies require personalized and guided interventions with clinical support which addresses the complexity of symptoms. Therefore, although computer-based self-help platforms have demonstrated positive outcomes, face-to-face CBT showed quicker and better reductions in abstinence rate and ED psychopathology in adults with EDs. [36].

As evidenced, while previous systematic reviews have compared guided to non-guided self-help platforms, there is a gap in research collating findings regarding the effectiveness of only computer-based psychoeducational and computer-based self-help platforms for people with EDs [11, 34, 37]. Therefore, this systematic review aims to evaluate the effectiveness of computer-based self-help platforms ± associated apps for several outcomes: (1) global ED symptoms, (2) ED-related behaviours, such as thin idealisation, body dissatisfaction, quality of life, depression, perseverative thinking, and resistance to change, and (3) preventing the onset of EDs. Researchers searched independently to increase the internal validity of the findings. It was performed per the Preferred Reporting Items for Systematic Reviews and Meta-Analyses (PRISMA) guidelines.

## Method

### Search strategy

A systematic search was conducted following the guidelines of the Preferred Reporting Items for Systematic Reviews and Meta-Analyses (PRISMA) (Page et al., 2021). The protocol for this systematic review has been registered on PROSPERO with registration number: CRD42042520866 and the protocol was published on JIMR journal. Four databases (Ovid MEDLINE I, Embase, Global Health, and APA PsychInfo) were used to identify relevant literature by the 31^st^ of May 2024. The search terms are depicted in Table 1. A manual search of references was conducted utilising Google Scholar to identify alternative literature. The search was conducted by AG and YYK and reviewed by EC.

**Table 1.**
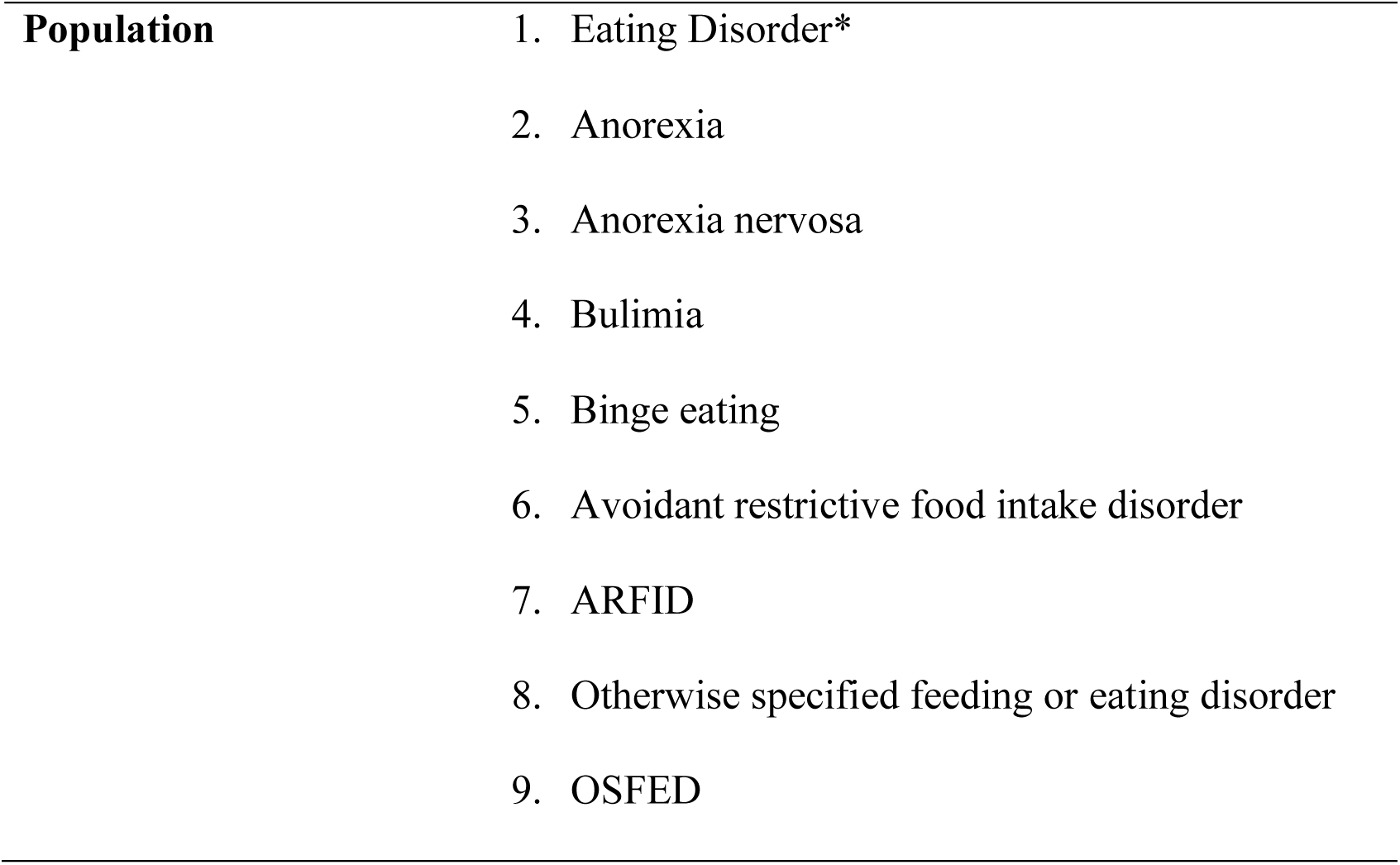
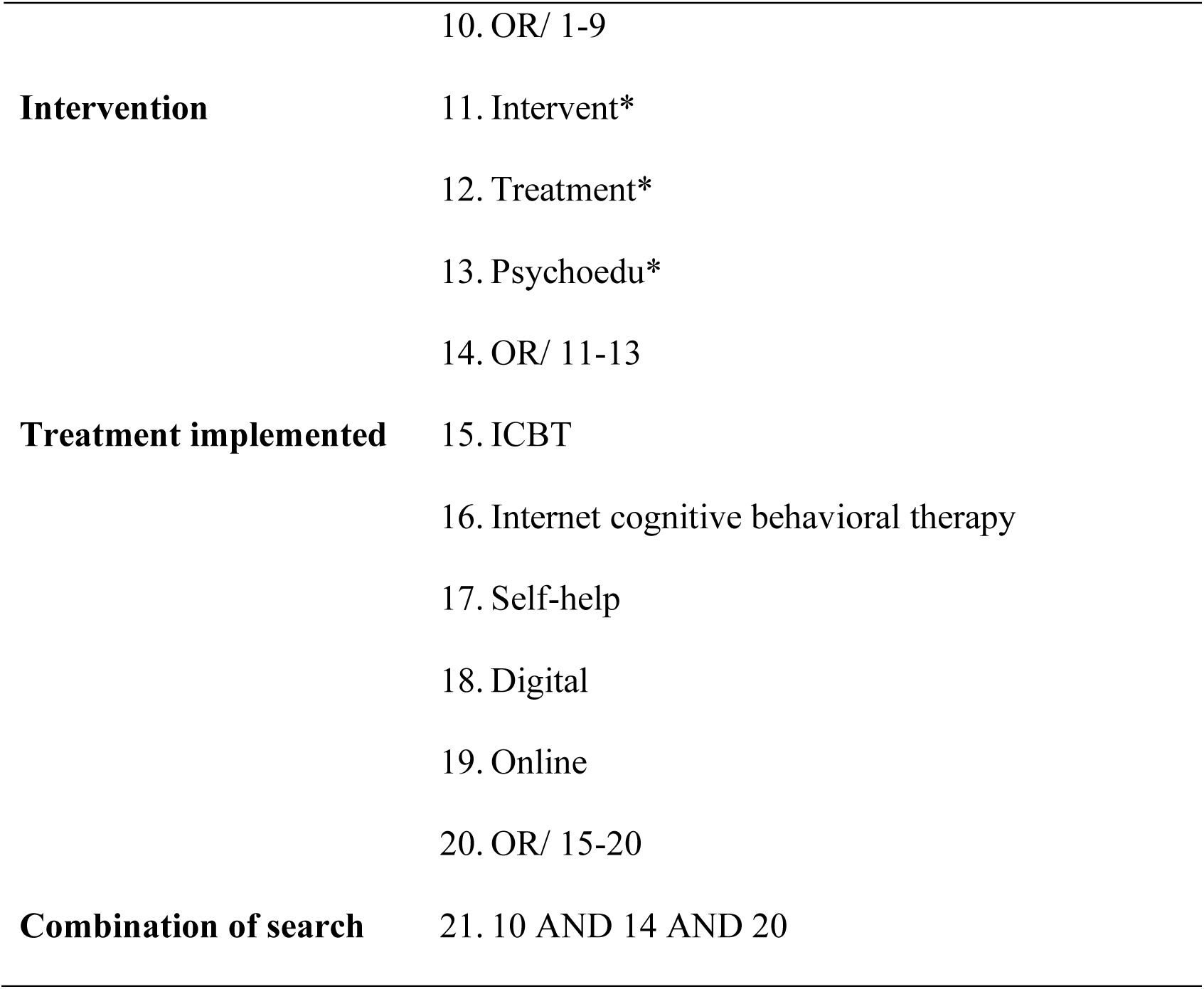
Example of search strategy on Ovid Medline.

| Population |  |
| --- | --- |
|  | 1. Eating Disorder* |
|  | 2. Anorexia |
|  | 3. Anorexia nervosa |
|  | 4. Bulimia |
|  | 5. Binge eating |
|  | 6. Avoidant restrictive food intake disorder |
|  | 7. ARFID |
|  | 8. Otherwise specified feeding or eating disorder |
|  | 9. OSFED |
|  | 10. OR/ 1-9 |
| **Intervention** | 11. Intervent* |
|  | 12. Treatment* |
|  | 13. Psychoedu* |
|  | 14. OR/ 11-13 |
| **Treatment implemented** | 15. ICBT |
|  | 16. Internet cognitive behavioral therapy |
|  | 17. Self-help |
|  | 18. Digital |
|  | 19. Online |
|  | 20. OR/ 15-20 |
| **Combination of search** | 21. 10 AND 14 AND 20 |

### Eligibility criteria

| **Inclusion** | **Exclusion** |
| --- | --- |
| Research with original data, including grey literature | Only guided self-help platforms with no self-help comparison group |
| Research evaluating the efficacy of computer-based psychoeducational self-help platforms for EDs | Only in-person interventions with no computer-based comparison group |
| People with ED diagnosis | Only in-person delivered interventions |
| People with ED symptoms | Self-help platforms for conditions other than EDs |
| People at risk of developing EDs | Systematic reviews |
| People from the general population without ED-related behaviors | Meta-analyses |
| Research which examined the pre- and post- computer-based intervention clinical outcome of ED symptoms and related behaviors | Posters |
| Research which examined the pre- and post- computer-based intervention associated mental health difficulties | Leaflets |
| Literature in English | Books |
|  | Reviews |
|  | Research which only reported physical outcomes |

### Screening strategy

After the search was completed, studies were exported to EndNote, and duplicates were removed. AG and YYK then initially screened articles independently by titles and abstracts. Following this, the authors screened the full text of the literature and collected research that abided by the inclusion and exclusion criteria. AG and YYK did not find any discrepancies. Any disagreements were resolved by discussion with the senior author, EC. The systematic review excluded literature that did not meet the inclusion criteria.

### Data extraction

AG and YYK independently completed data extraction. The key aspects were recorded on an Excel table based on the following variables: the primary author’s name and published date; participants profile, baseline sample number, female gender percentage and mean age; follow-up times and sample size; outcome measure; intervention and comparison group; key findings; and overall risk of bias. AG and YYK inputted the data independently for each included study to assess the study’s eligibility. There were no discrepancies during data extraction. EC would have been consulted for any uncertainty or unresolved disagreements.

The included studies were separated into three categories based on the participant populations and the implemented intervention. Based on the stepped and THRIVE approach, studies included in the primary prevention were involved participants from the general population. Studies included in the secondary prevention involved participants who were at risk of developing EDs and studies in the tertiary prevention included participants meeting the clinical ED threshold. The authors categorised thin idealisation, body image, body shape concerns, and weight concerns as ED-related behaviours.

### Quality assessment

The Grading of Recommendations Assessment, Development and Evaluation (GRADE) method was used to analyse the quality of the studies. The GRADE method was performed to analyse the risk of bias, inconsistency, indirectness, imprecision, and publication bias ranging from ‘no serious inconsistency’ to ‘very serious’. Additionally, the revised Cochrane risk-of-bias assessment (RoB 2) was utilised to assess the risk of bias of the randomised controlled trials (RCTs) using the Cochrane Review Manager software. All included literature were RCTs, and no additional research methods were detected. The five domains of potential risk-of-bias explored were randomisation, divergence from the intended intervention, analysis of missing data, measurement of outcomes, and selection of reported results. AG and YYK independently reviewed each included study and determined levels of potential risk-of-bias for each domain, ranging from ‘low’ to ‘high’. The reporting of the study adhered to the PRISMA 2020 guidelines.

### Strategy for data synthesis

The systematic review findings were summarised in an Excel table and then narratively synthesised and analysed based on Popay et al.’s (2006) guidelines. Included studies were analysed for clinical effectiveness and ED related behaviors, and ED prevention). Clinical effectiveness was reported in terms of the computer-based self-help platforms ± associated apps’ effectiveness in decreasing the ED symptoms and psychopathologies. ED related behaviors were reported in terms of the self-help platforms ± associated apps’ effectiveness in reducing ED related mental health difficulties (i.e. perseverative thinking, body dissatisfaction, thin idealization, fear of becoming fat, preoccupation of food and weight, motivation to change their weight, self-esteem, depression, and quality of life). ED prevention was reported in terms of the ability of the computer-based self-help platforms ± associated apps to reduce the onset of ED eating behaviors.

Most of the studies had a high amount of missing data, so the final data gathered from participants who completed the study was assessed. There were variances in the population target of the included studies, thus, the authors split the findings into three sections according to the population: primary, secondary, and tertiary prevention. Primary prevention was used for research in generally healthy populations, secondary prevention was used for research conducted with participants who were at risk of developing EDs, lastly, tertiary prevention was used for research in participants meeting the clinical threshold of EDs. These domains were established to make the effect of the treatment in different population groups comparable.

## Results

### Study Selection

The review identified a total of 4687 studies. Following the removal of duplicates (*n=* 2007), 2680 studies remained. After screening of titles and abstracts, 2545 studies were ineligible based on the exclusion criteria. 146 studies were screened by full text. 134 studies were excluded based on the exclusion criteria and fourteen studies were subject to narrative synthesis and included in this systematic review. Most studies were excluded due to having a guided feature and no self-help comparison group or due to the self-help intervention not being virtually delivered, but rather delivered via booklets. Figure 1 depicts the PRISMA flow-chart of the selection process. Overall, fourteen studies were eligible for narrative synthesis.

**Figure 1.**
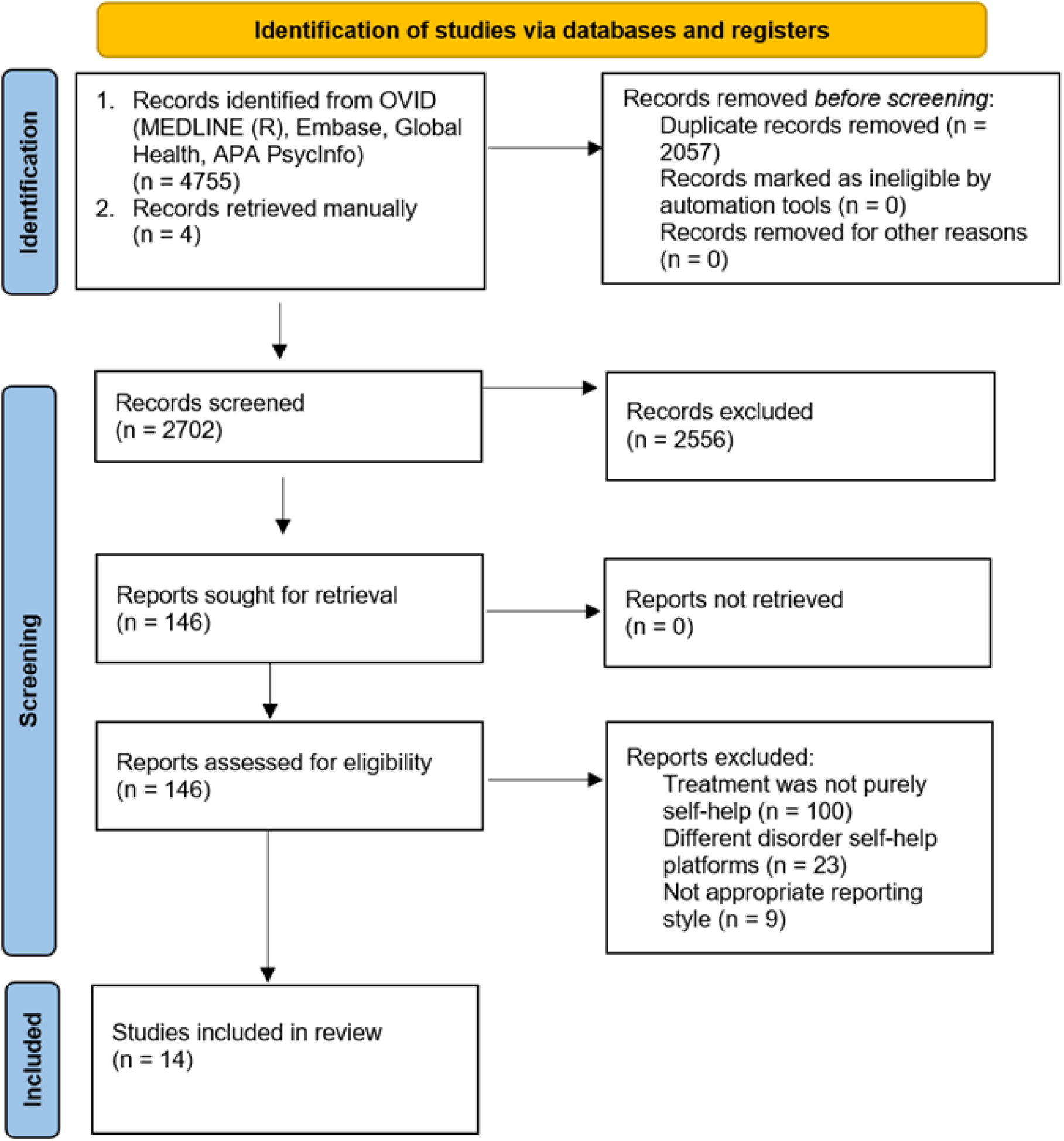
PRISMA flow chart of study selection.

### Study Characteristics

The included sample size ranged from 89 to 680 participants (*n=* 4195). Participants were recruited from the general public, from individuals at risk of developing EDs, or from those who met the clinical criteria for or were diagnosed with an ED. The mean age of participants was 22.9 years, ranging from 15 to 50 years old. 98.8% of participants were female. All studies included were RCTs. Studies took place in four continents, Europe (*n* = 6), North America (*n=* 4), Asia (*n* = 1), and Oceania (*n=* 3). All studies utilised self-report measures and employed questionnaires which measured the clinical effectiveness and mental health outcomes of the intervention, related to ED symptoms and ED related behaviors. The characteristics of the included studies are depicted in Table 2.

**Table 2.**
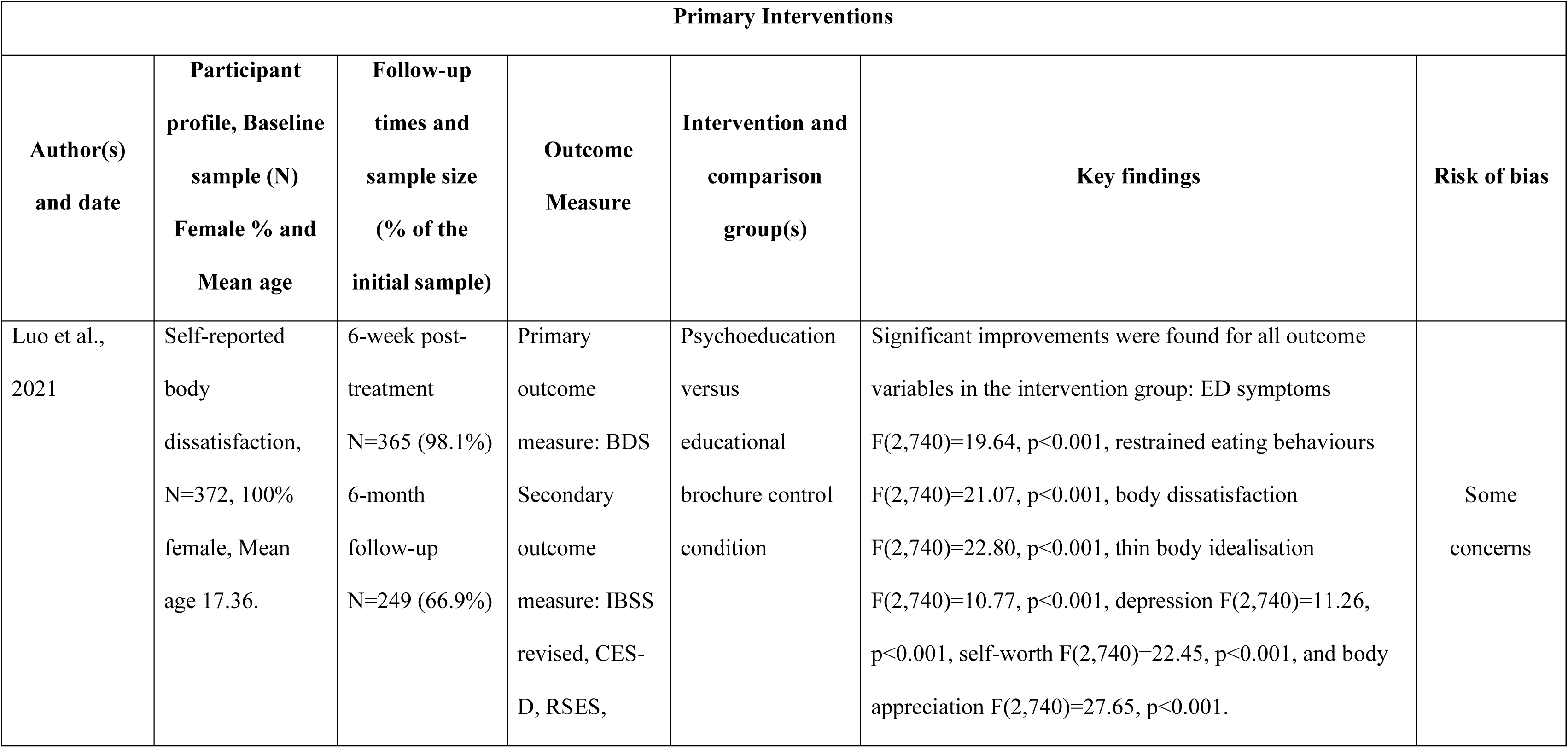
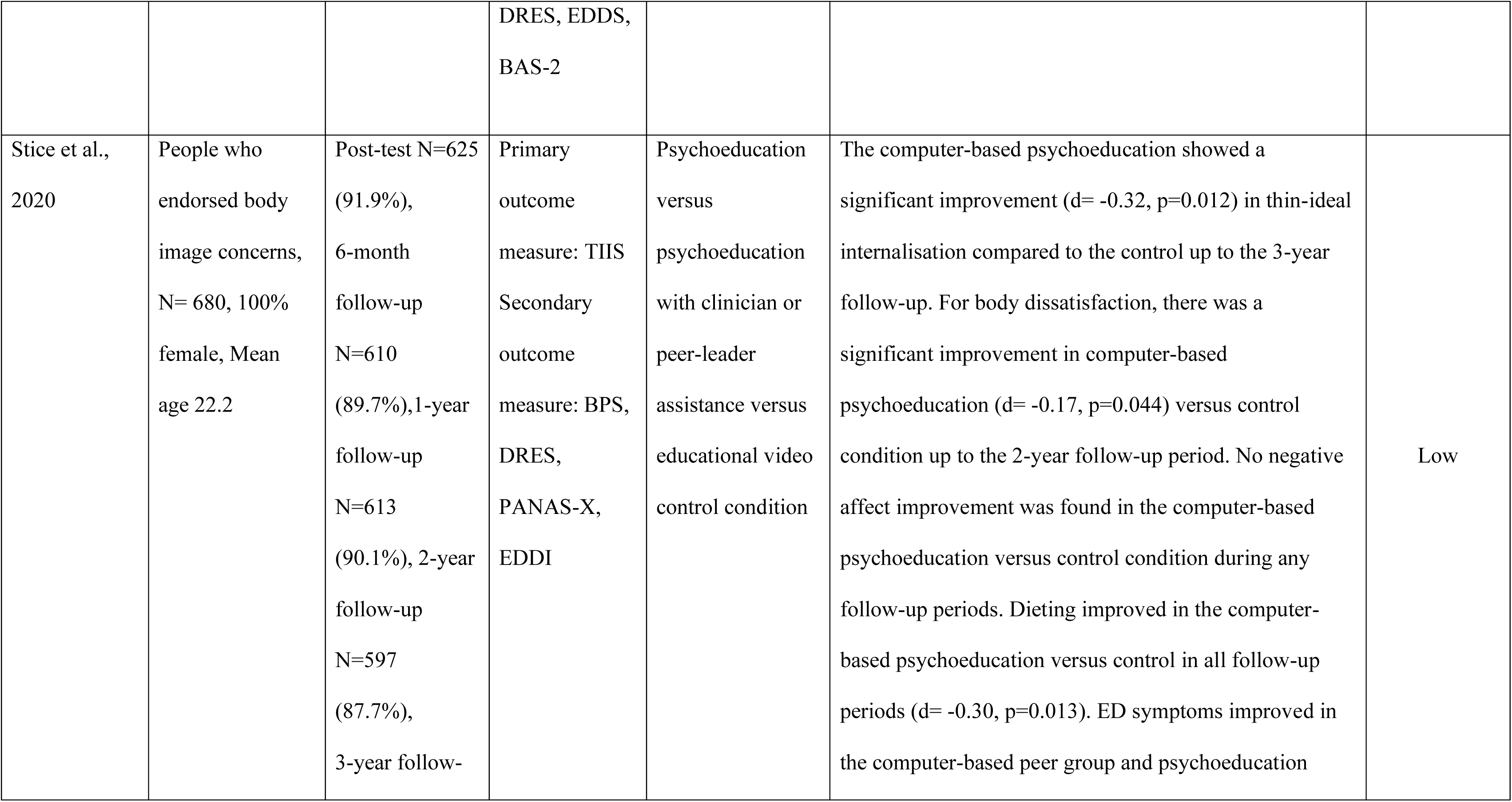
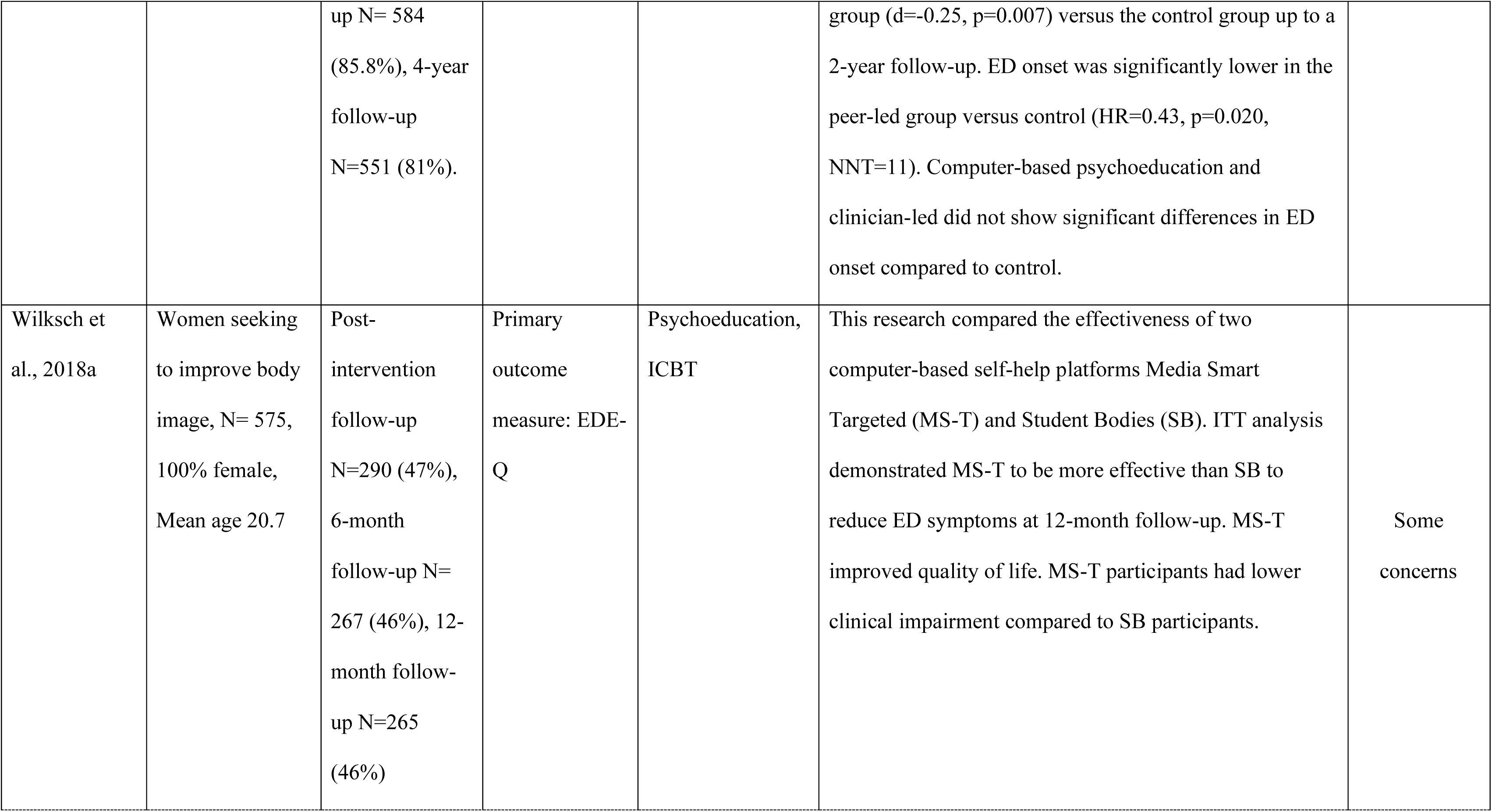
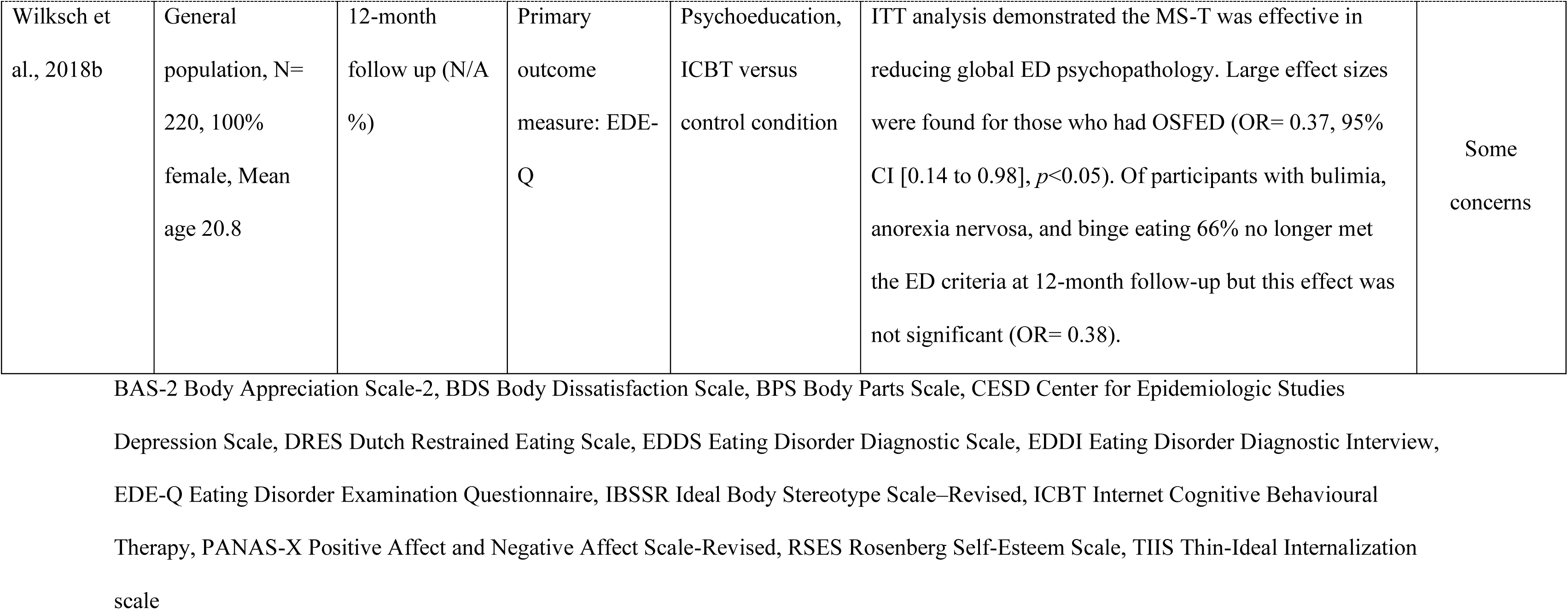
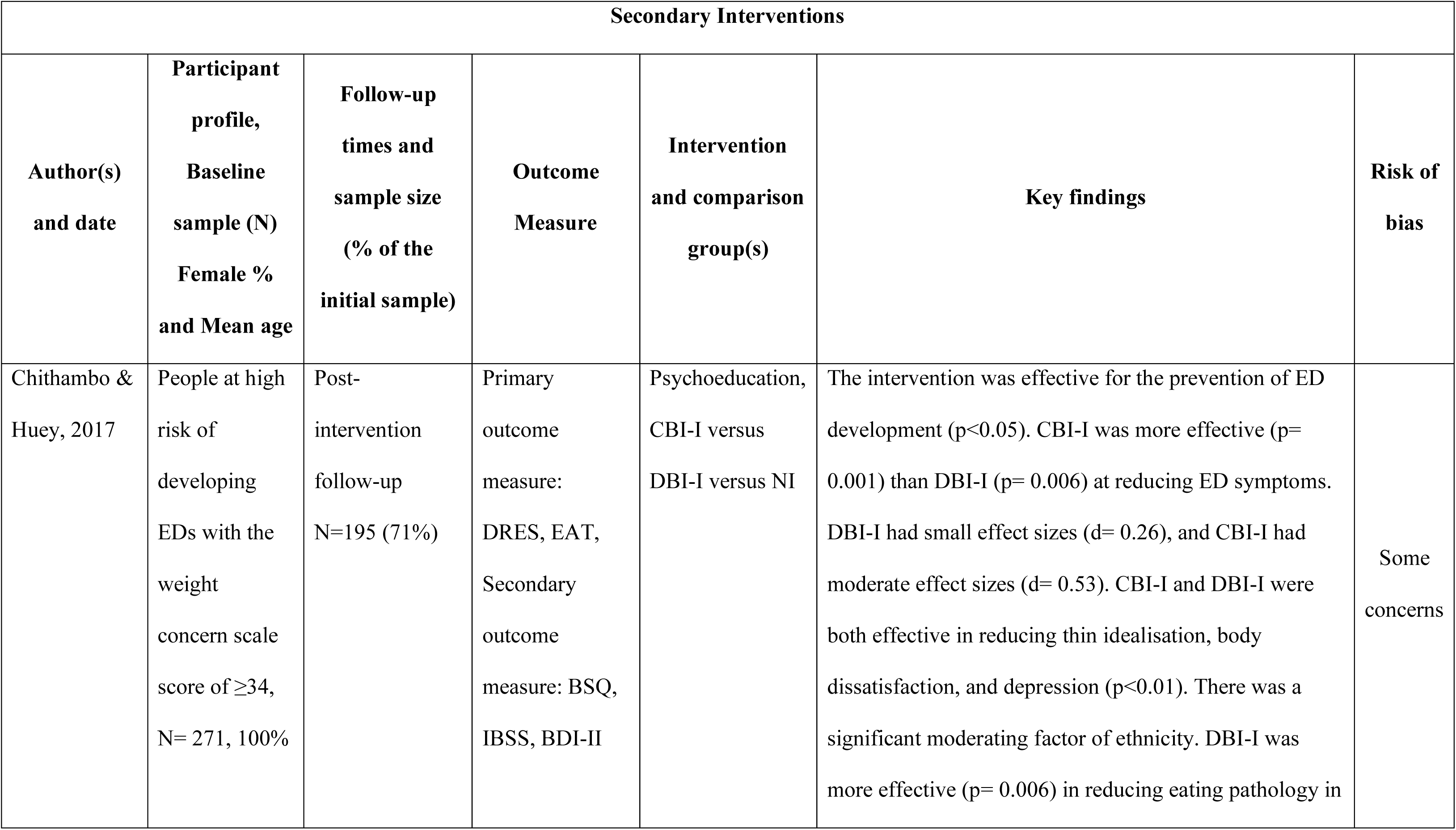
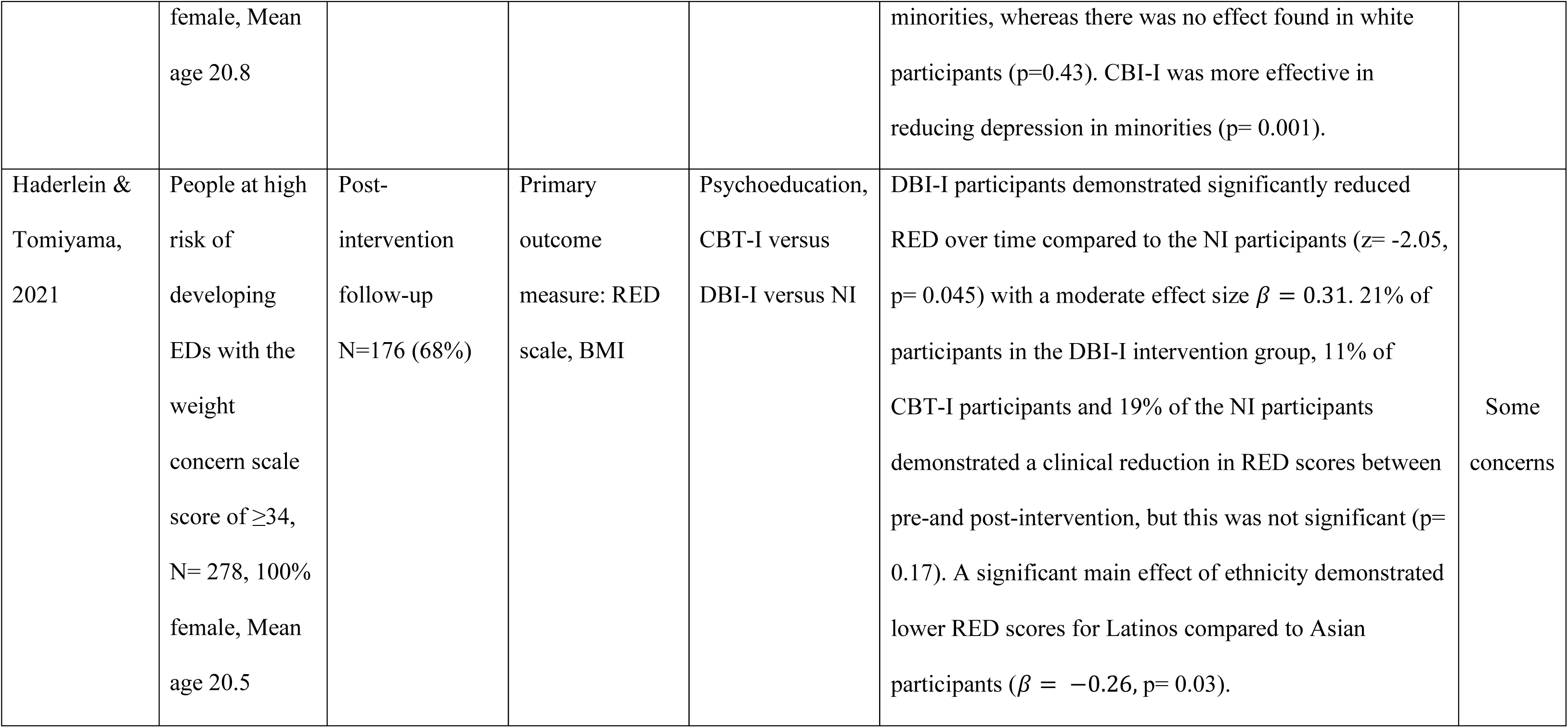
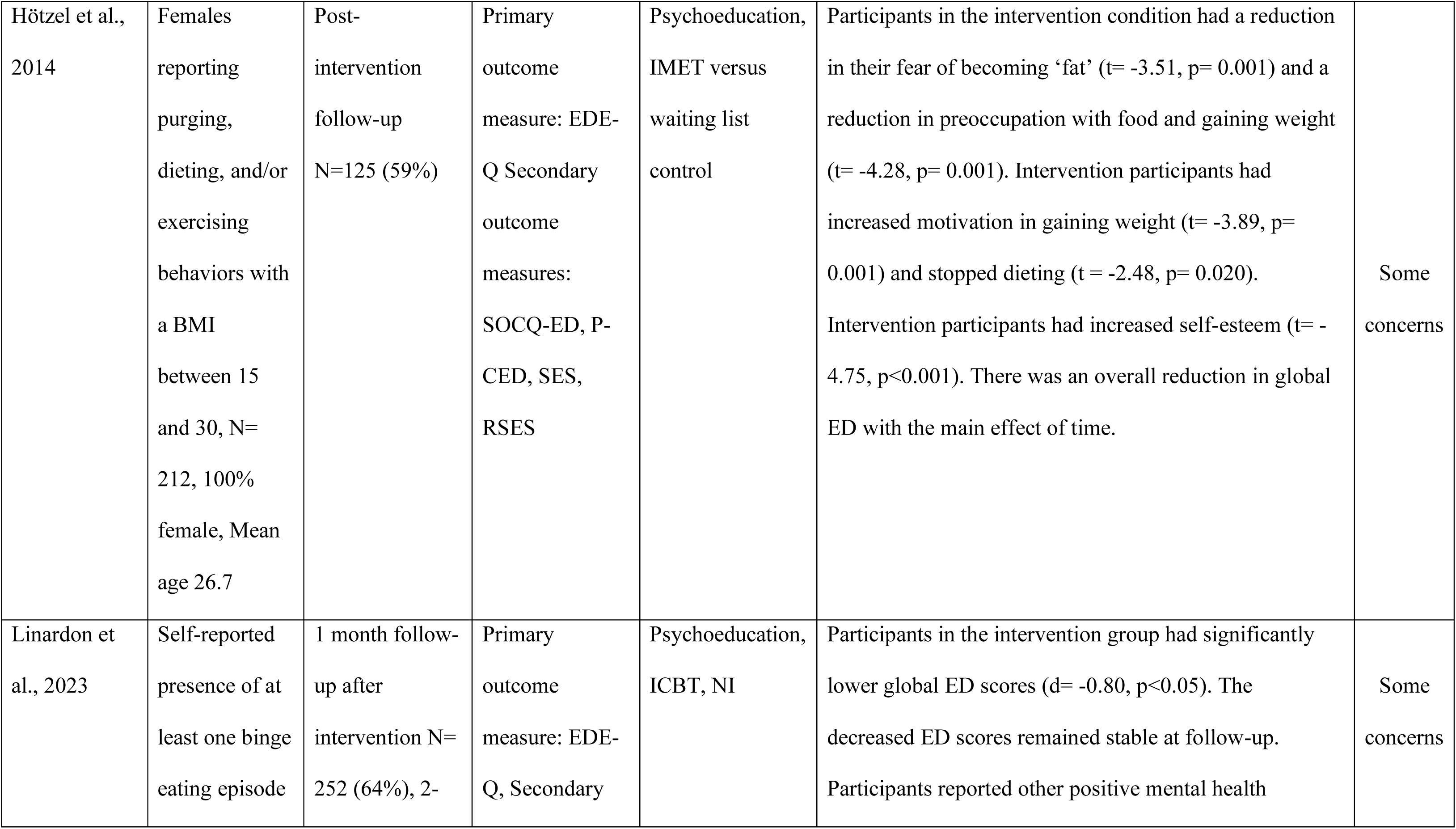
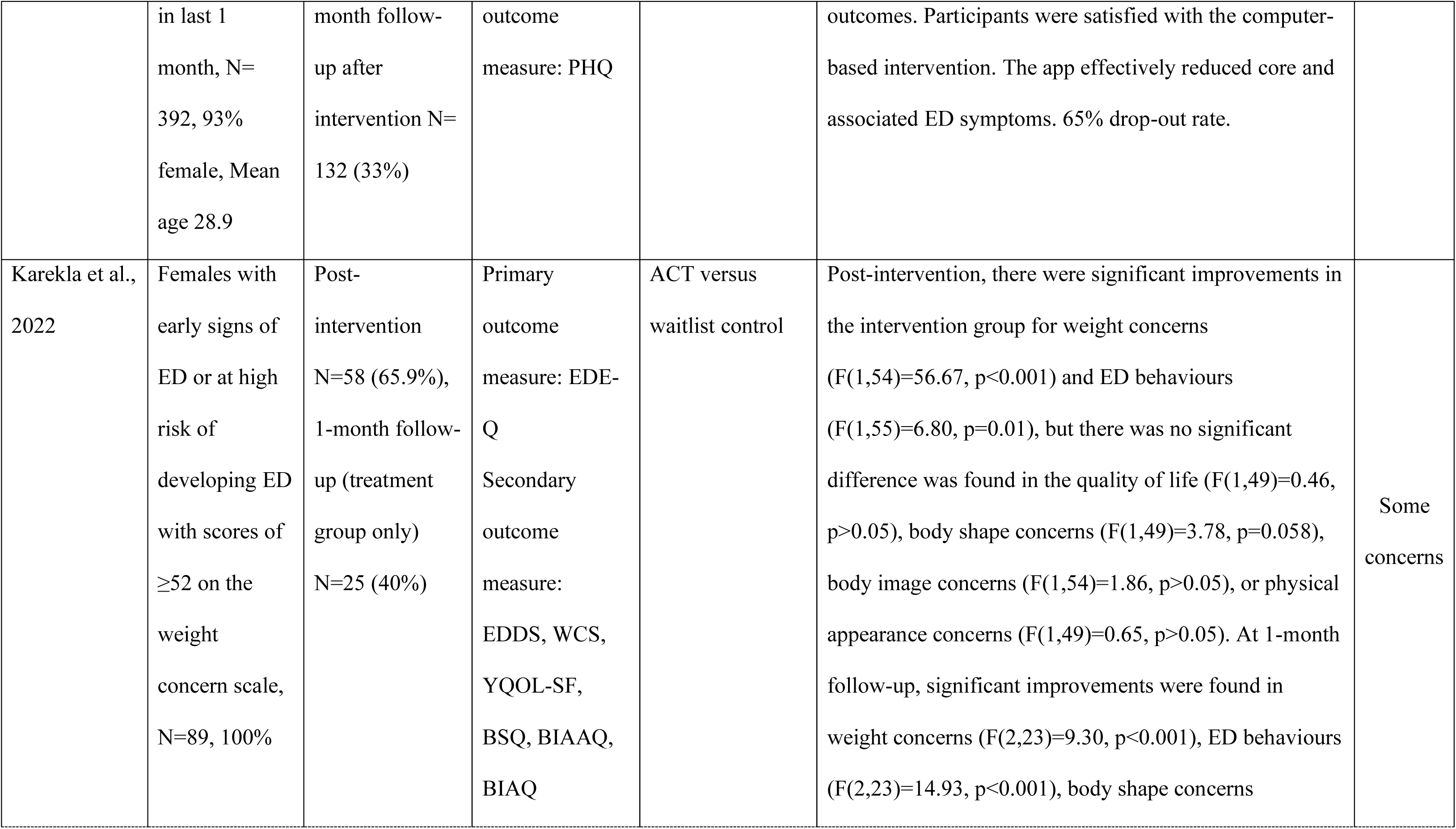
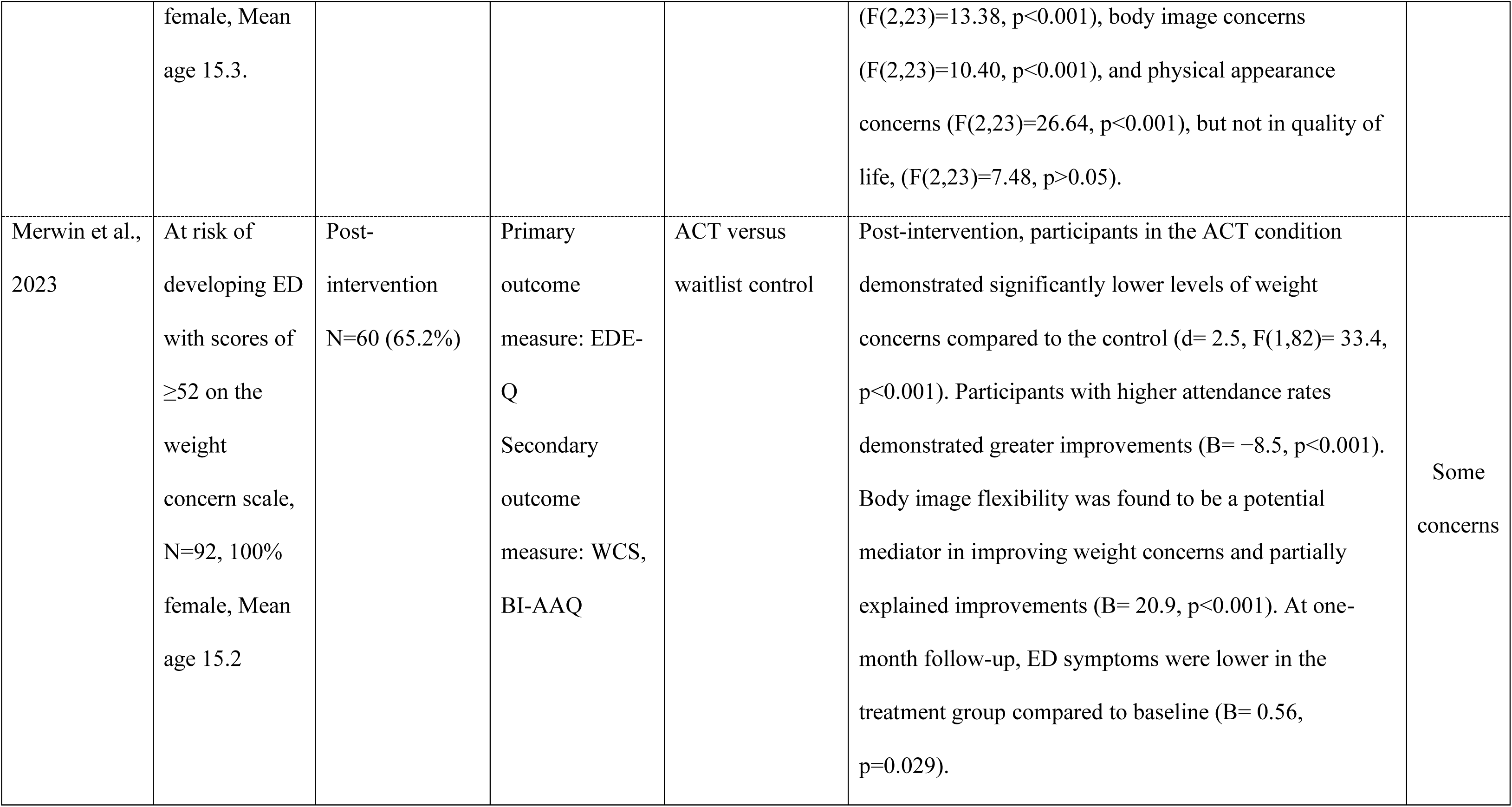
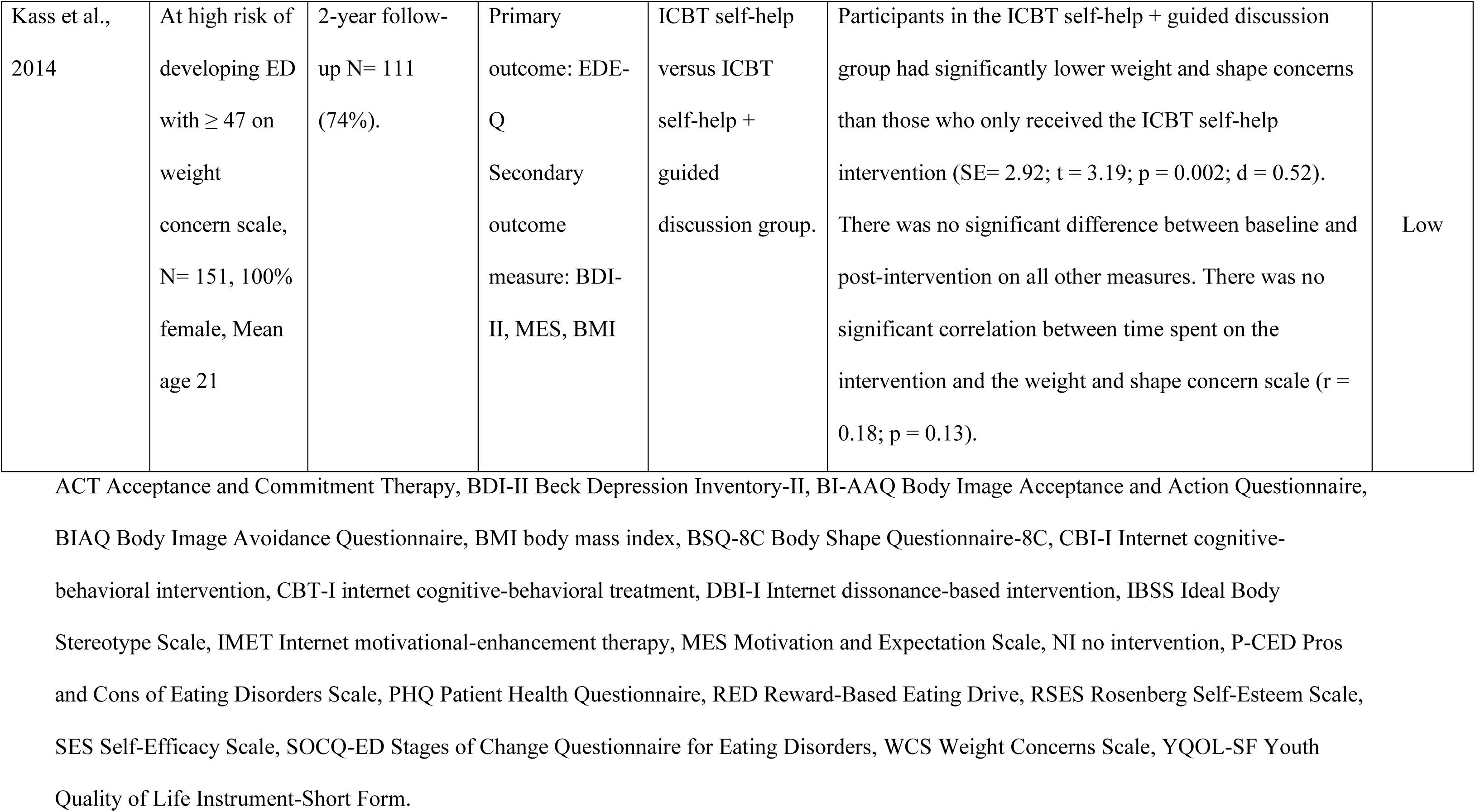
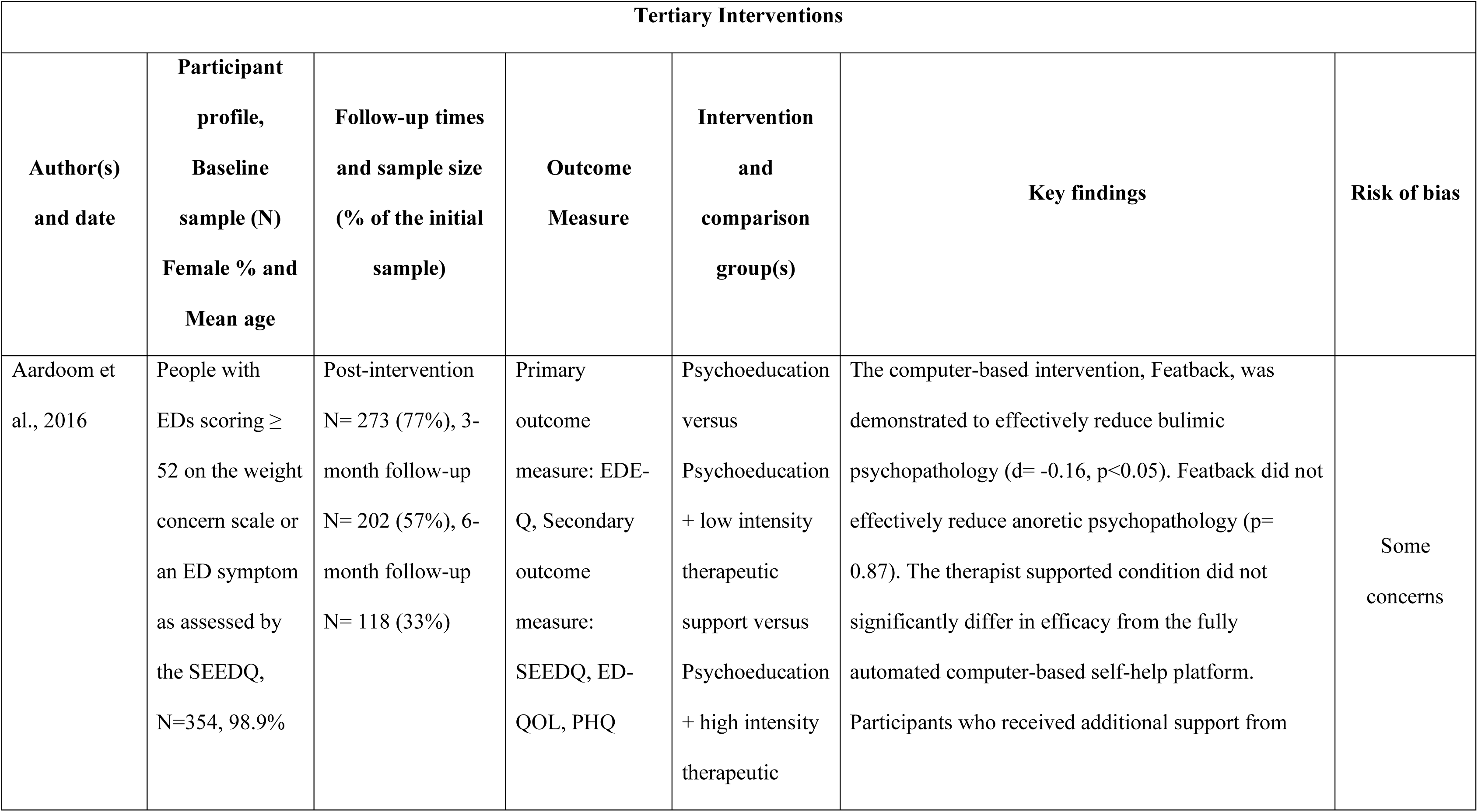
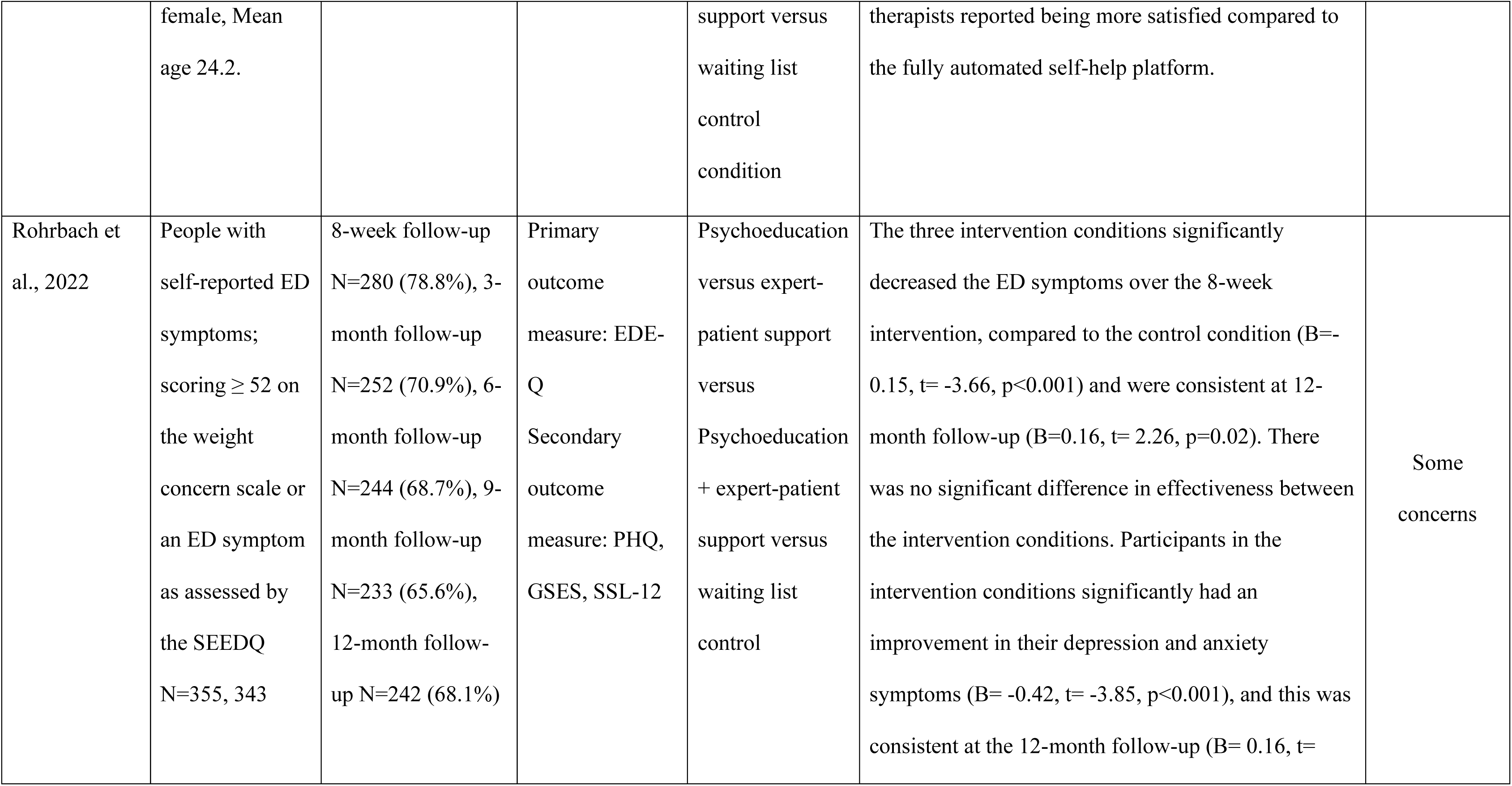
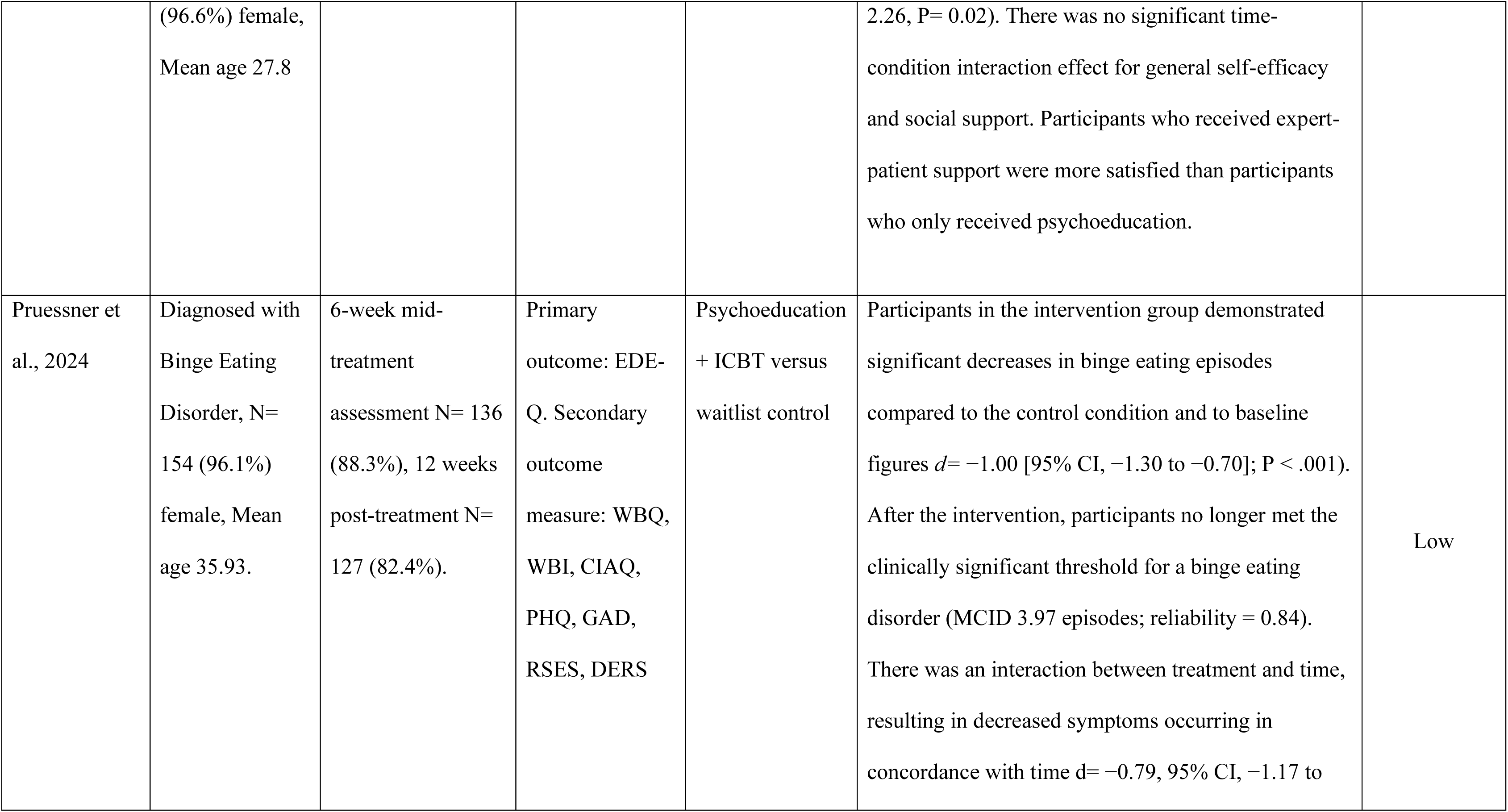
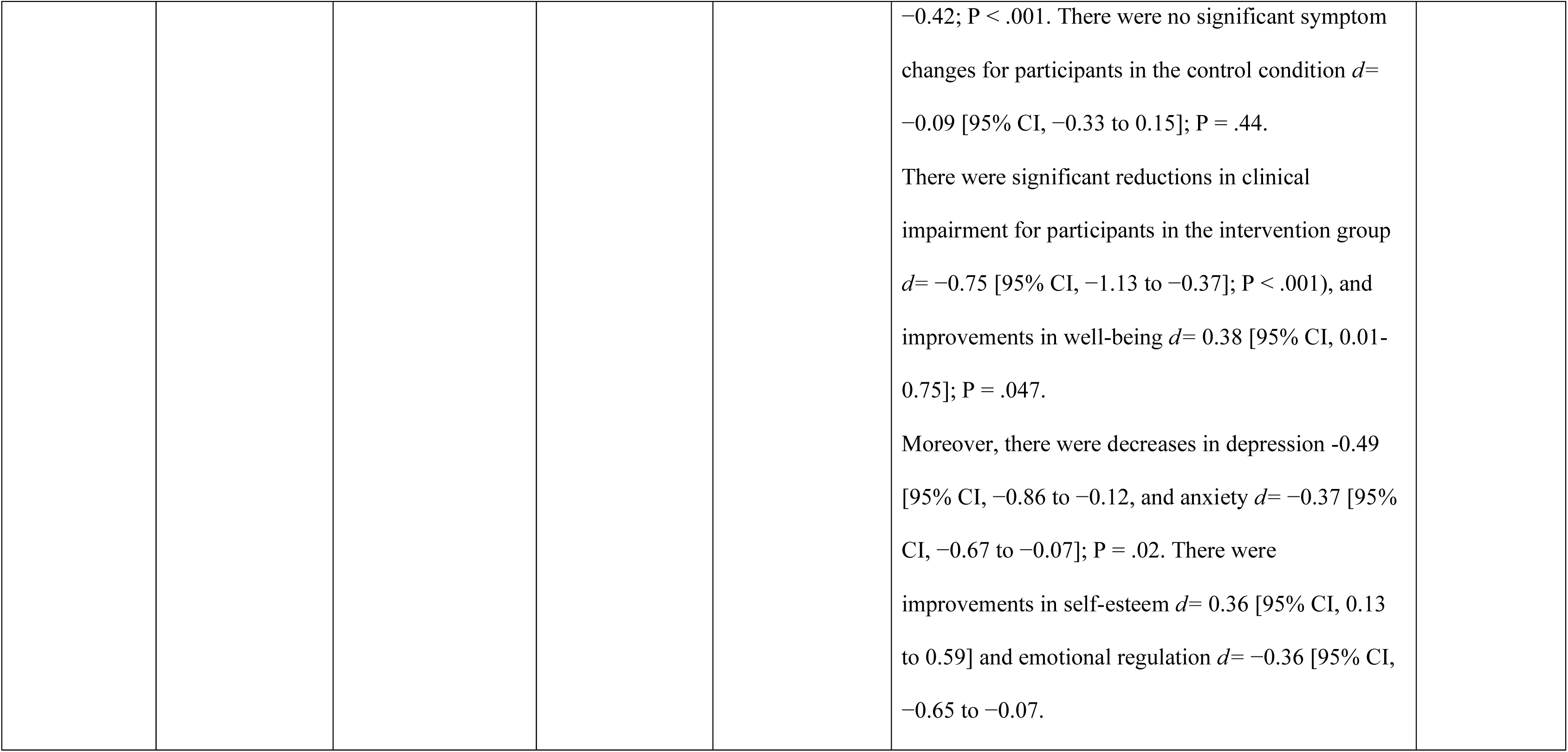
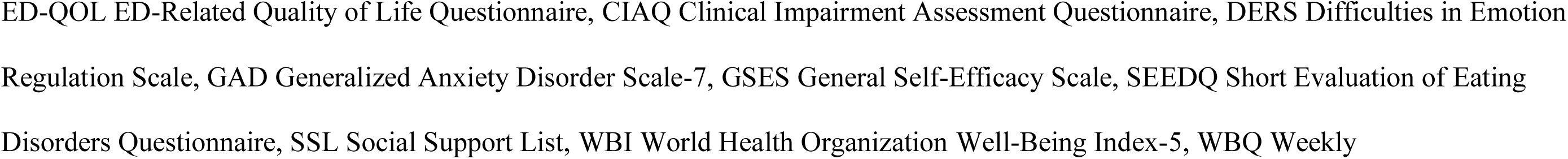
Characteristics of included studies.

### Quality assessments and Cochrane risk-of-bias analysis for included studies

The risk of bias assessment for each study is presented in Figure 2. Only one study showed an overall low risk of bias [14]. Of the 14 included RCTs, missing outcome data was the most common cause for bias, found in *n*=9 (64%) studies, with low missing data found in the rest of the studies [14, 33, 38]. Only three studies showed measurement bias, making it the least common cause of bias [38–40]. All studies found no risk of bias due to deviations from the intended intervention. Overall, the risk of bias among the studies was mainly categorised as some concerns. The percentage of the risk of bias is presented in Figure 3. Most studies are of moderate quality. The GRADE profile of the studies is shown in Table 3.

**Figure 2.**
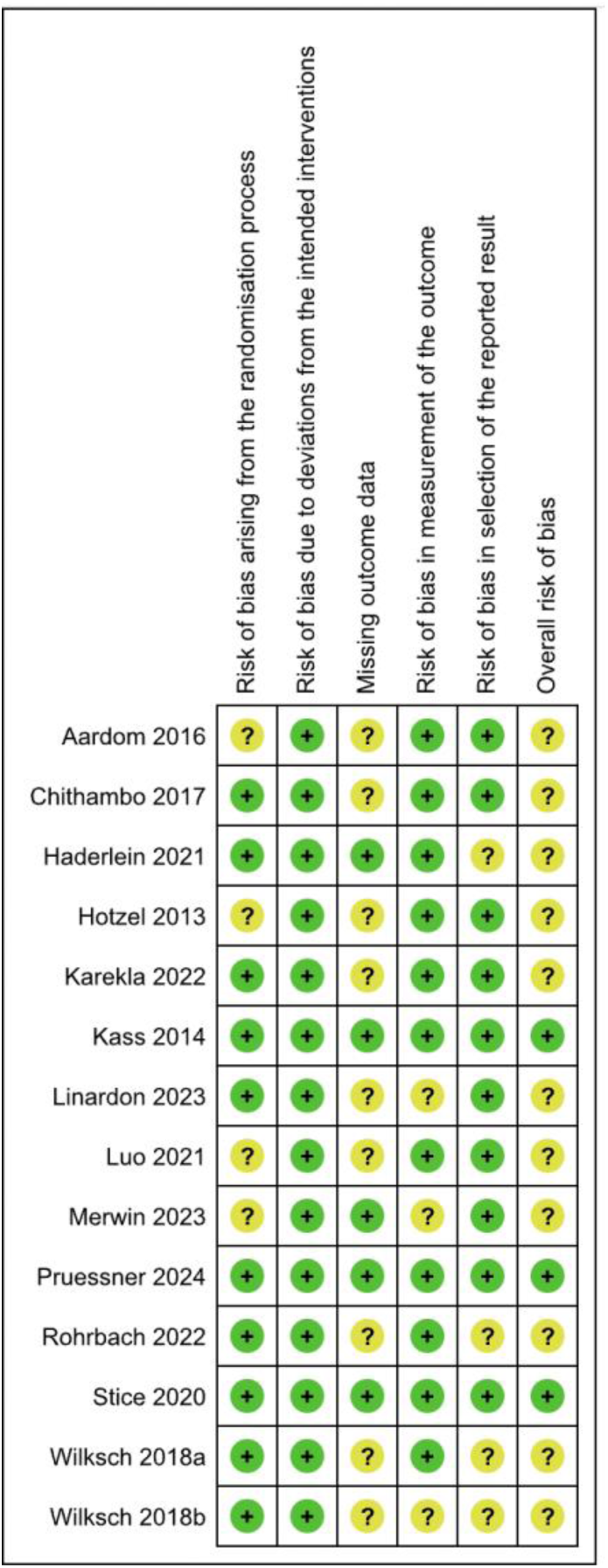
Risk of bias table.

**Figure 3.**
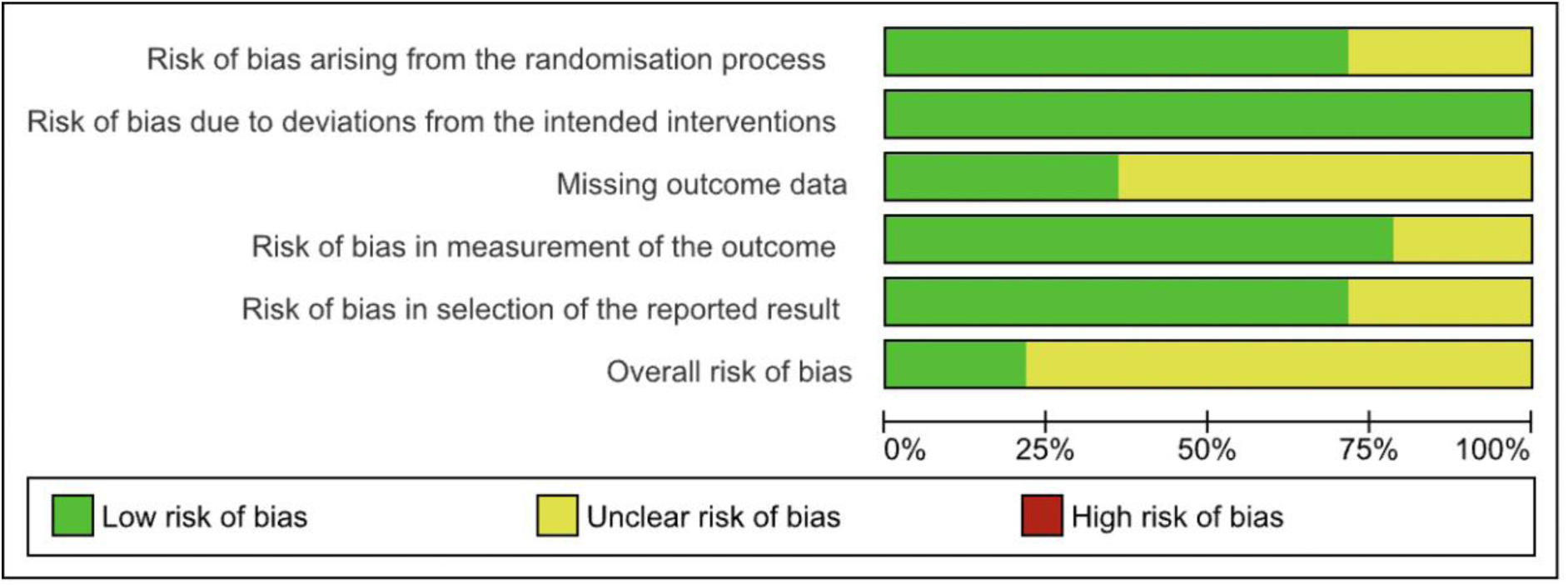
Risk of bias percentage table.

### Effectiveness of computer-based self-help platforms ± associated apps

This systematic review included studies conducted with the general population (*n*= 4) [14, 39, 41, 42], with participants at risk of developing EDs (*n=* 7) [30, 31, 33, 38, 40, 43, 44], and participants with ED symptoms or EDs (*n=*3) [32, 45, 46] Most studies (*n=*10) utilised the EDE-Q to measure changes in ED psychopathology, which has been found to be highly sensitive for ED detection, 0.83 and a specificity of 0.96 (Cronbach’s α=0.85 to 0.93, reliability r=0.68 to 0.74) [47, 48]. Other ED psychopathology tools included DRES (α = 0.95, r=0.82), IBSS (α = 0.73, r=0.80), and BSQ (α = 0.96, r=0.88).

All included studies demonstrated that participants in the intervention groups demonstrated significant improvements in ED symptoms and ED-related behaviors compared to the control conditions. Additionally, studies that utilised unguided computer-based self-help platforms ± associated app interventions as a primary prevention tool demonstrated them to be effective. Moreover, the studies found that the positive effects were sustained at different follow-up times, which were up to a 4-year follow-up.

### Comparing modalities

Most studies (*n*= 11) utilised an element of psychoeducation within their intervention. On top of psychoeducation, the studies implemented ICBT techniques to address cognitive and behavioural changes in participants. The included studies which utilised psychoeducation in combination with ICBT demonstrated significantly better effectiveness compared to psychoeducation alone and psychoeducation with other modalities. Moreover, acceptance and commitment therapy (ACT) and motivational enhancement therapy (MET) were also found to reduce ED psychopathologies effectively.

Two studies compared ICBT to IDBI combined with psychoeducation. The findings are contradictory, as one of the studies suggests that ICBT is more effective than IDBI in reducing global ED psychopathologies [31]. However, the alternative study demonstrated that IDBI was more effective than ICBT in reducing reward-eating behaviours [33]. The findings demonstrated that ICBT was overall more effective compared to IDBI; however, there was a main effect of ethnicity. The results show that ethnicity has a significant impact on the effectiveness of the intervention. IDBI was demonstrated to be more effective in minority participants compared to white participants. The main effect of ethnicity suggests that different interventions may be more effective for the global majority groups versus white participants. Table 4 provides an overview of the comparison between the modalities.

**Table 4.**
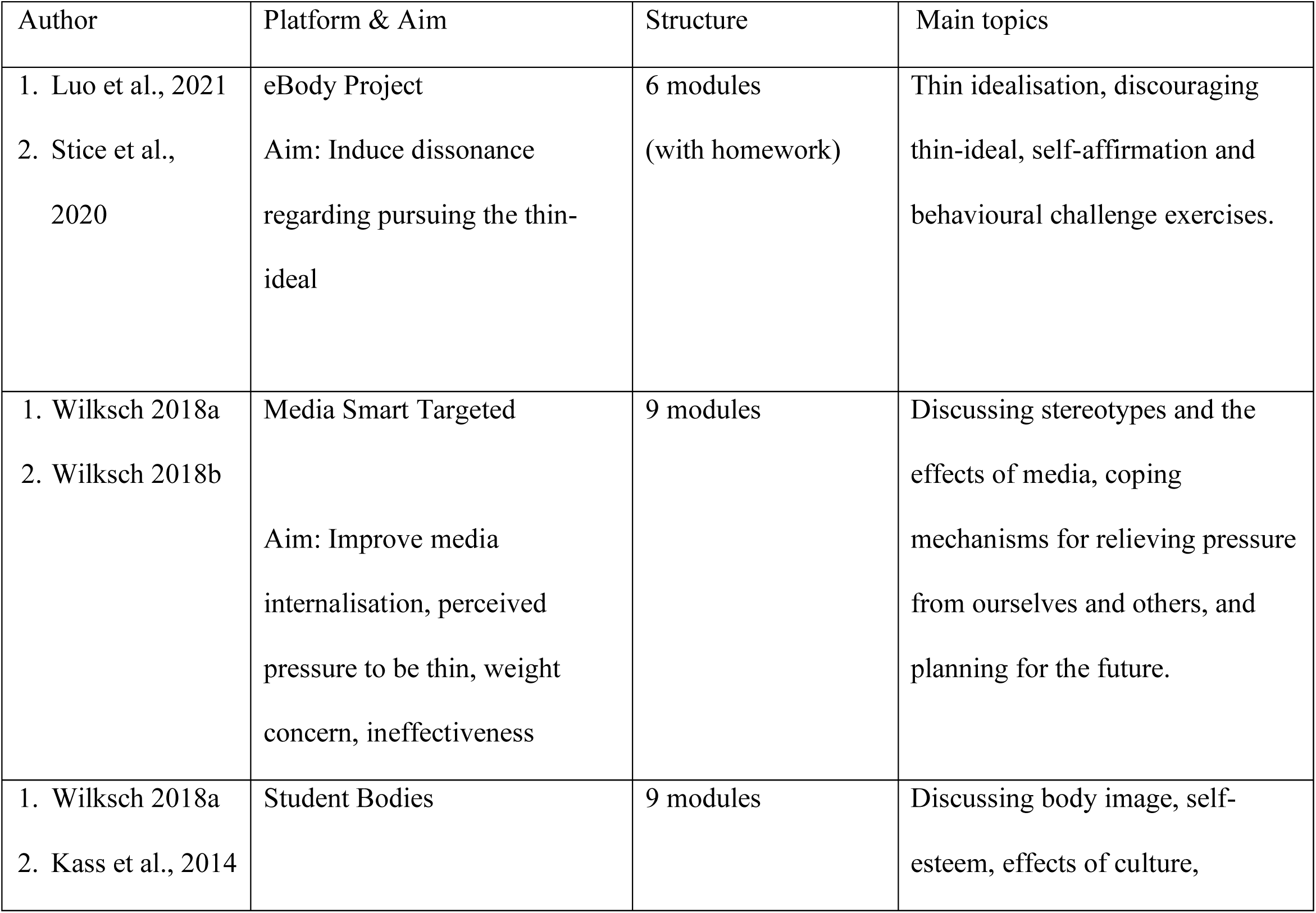
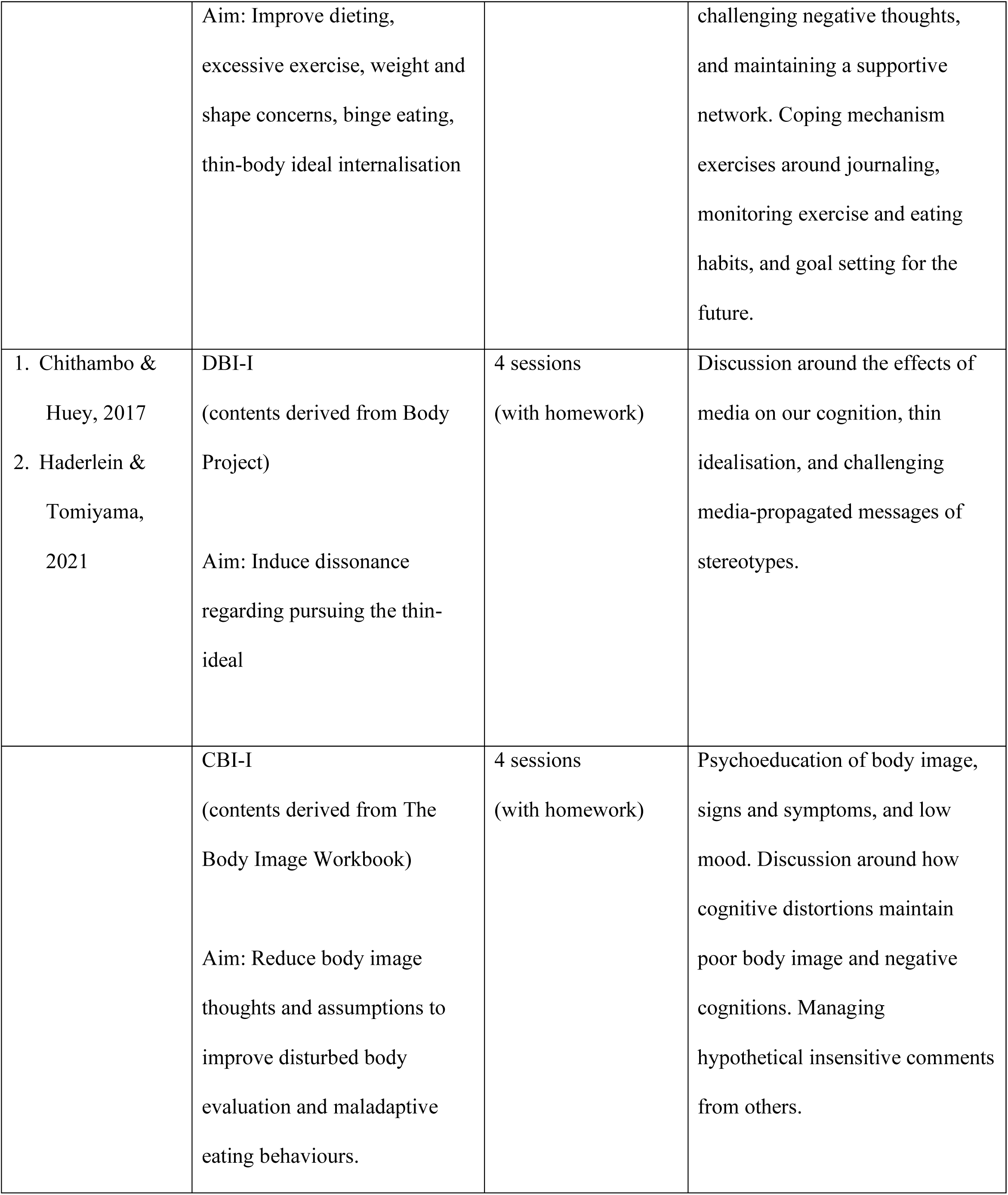
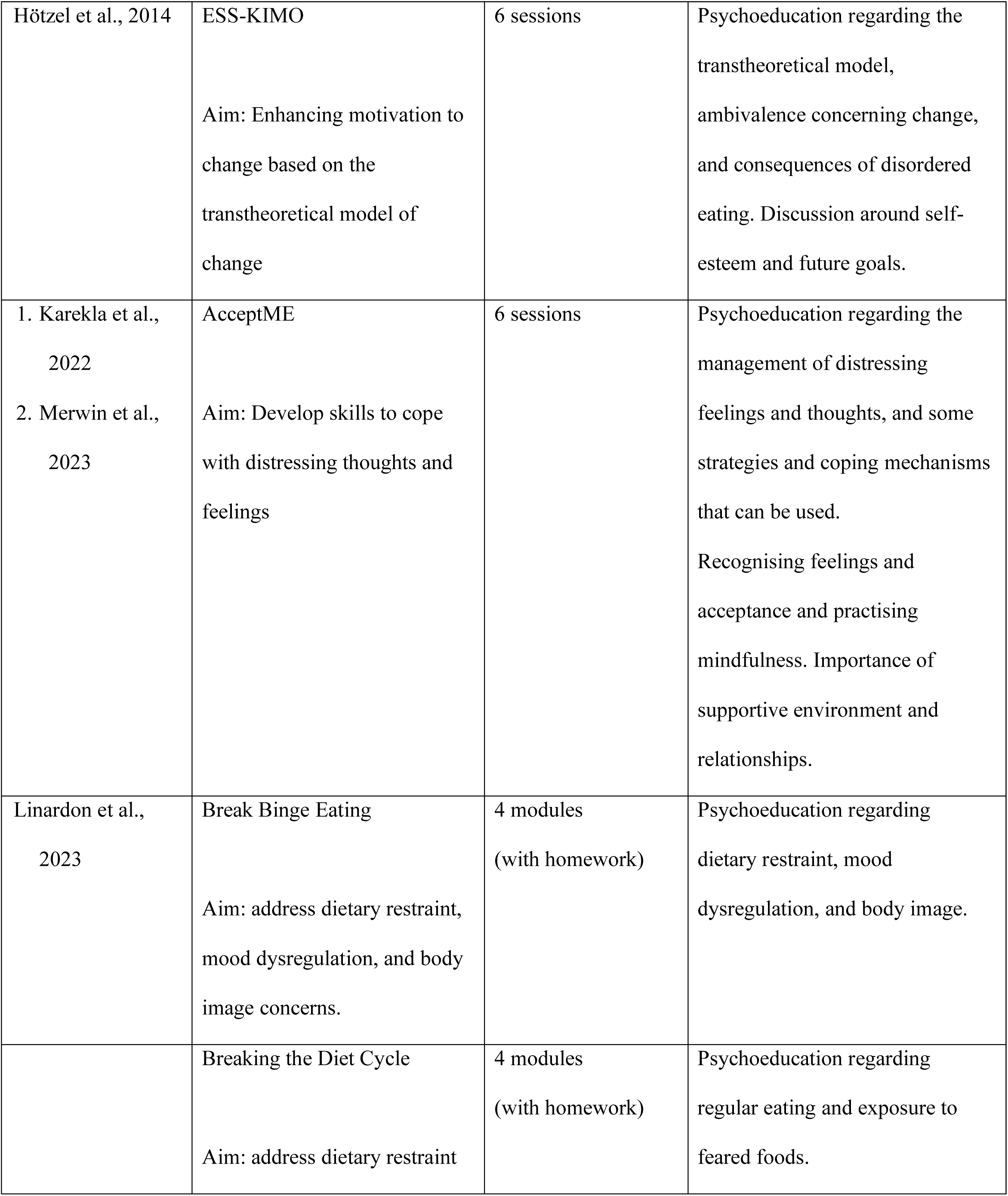
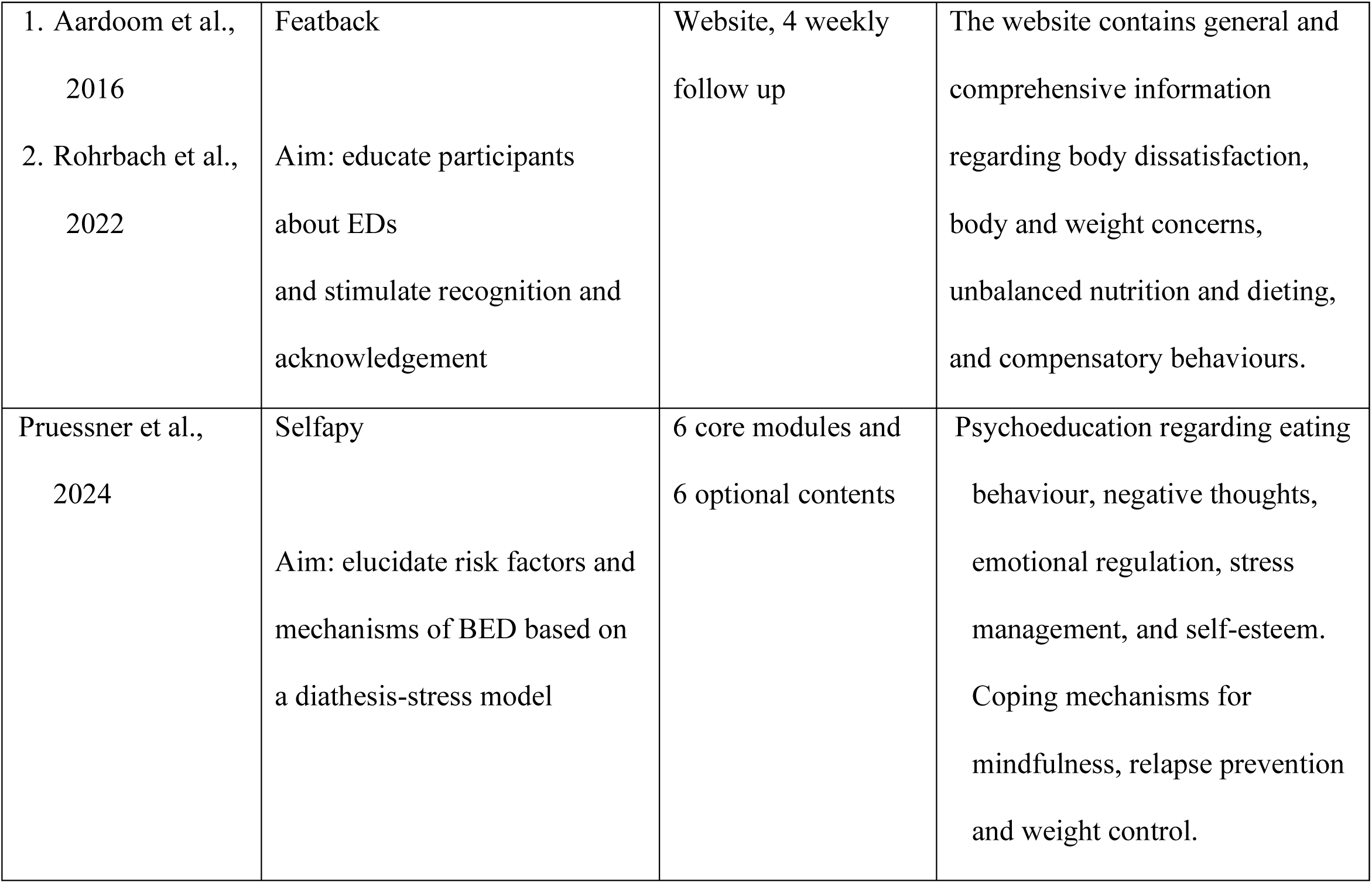
Comparison between modalities.

### Guided versus Unguided modalities

Some included studies (*n*=4) included a guided group alongside the unguided modality [14, 30, 32, 46]. While unguided modalities showed significant improvements compared to control conditions, participants in guided interventions experienced even greater reductions in symptomatology. Additionally, participants in the guided group conditions were significantly more satisfied than participants in the unguided conditions [32, 46]. Lastly, Stice et al. (2020) demonstrated significantly lower ED onset rates for participants in the peer-led groups compared to unguided and clinician-guided psychoeducation conditions.

### ED-related behaviours

Alongside global ED psychopathology, all studies measured changes in ED-related behaviours and comorbidities. All questionnaires used had good validity, with reliability scores of YQOL-SF (α = 0.80), BDI-II (α = 0.90), TIIS (α = 0.75, r=0.56), and BDS (α = 0.94, r=0.90). DBI-I had a smaller effect size (d=0.26) than CBI-I (d=0.53) in reducing ED-related behaviours. The findings demonstrated significant improvements in ED-related behaviours post-intervention compared to the control condition or baseline levels.

Furthermore, ICBT was effective in improving self-esteem (d=0.36) and emotional regulation (d=-0.36) in patients with EDs. The findings demonstrated the improvements to be sustained at 1-month follow-up. Contrastingly, one study which utilised ACT found no significant improvement in ED-related behaviours. Interestingly, while ICBT and IDBI were both found to be effective in reducing depressive symptoms, ICBT was found to be more effective in minority groups compared to white participants [31].

## Discussion

This systematic review found that self-help platforms are effective in reducing global ED behaviours and ED-related behaviours. Moreover, computer-based self-help platforms ± associated apps for EDs are perceived positively by users as they reduce the barriers to getting support. In line with the THRIVE framework, the self-help platforms were most effective when they were utilised as preventative measures [24]. Computer-based programs have the potential to give the public access to evidence-based psychoeducational tools and resources [24]. This systematic review collates the clinical effectiveness of a range of self-help platforms, despite different outcome measures being used. Self-help platforms show a promising avenue for the management of ED symptoms, particularly when looking at a 6-month follow-up time.

Traditionally, especially pre-COVID, many therapeutic interventions were delivered face-to-face. For EDs, face-to-face family-based treatment for EDs and CBT-ED were the most commonly utilised therapeutic interventions supported by numerous evidence-based clinical effectiveness studies [49–52]. After COVID-19, interventions that were previously delivered face-to-face transitioned to computer-based self-help platforms to educate and provide resources aimed at effectively preventing and treating ED symptoms and related behaviours. While conducting this systematic review, the authors found that computer-based interventions follow similar principles to the traditional face-to-face principles. Most computer-based self-help platforms are purely psychoeducational, CBT format (ICBT), or DBT format (IDBI). While face-to-face interventions require an in-person meeting between clinicians and patients, computer-based interventions can be completed without assistance and overcome these barriers while maintaining the treatment effectiveness.

As shown by the results in this systematic review, of these computer-based self-help platforms, 7 platforms were CBT-based, 2 of which looked at primary prevention, 4 looked at secondary prevention, and 1 looked at tertiary prevention. Similar to face-to-face CBT, ICBT platforms showed positive clinical effectiveness, particularly in patients with BED and BN. When evaluating these results, it’s important to note that various outcome measures were employed to assess clinical effectiveness, including the EDE-Q, DRES, EAT, IBSS, and BDI-II, all of which demonstrated promising findings. Across the board, 5 findings showed reductions in global EDE-Q scores; 2 findings showed a reduction in abnormal eating behaviours as measured by RED, DRES, and EAT; 2 findings showed a reduction in distorted body image and pursuit of thinness as measured by BSQ and IBSS; and 2 showed a reduction in depression, measured by BDI-II. The findings of this systematic review align with the wider literature.

When compared to face-to-face CBT, the findings are similar. A meta-analysis containing 79 studies [49] demonstrated that therapist-led CBT reduced short-term remission and binge or purge frequency in BN and BED compared to waitlisted conditions. A meta-analysis of 16 studies exploring ICBT effectiveness found that ICBT was effective in preventing ED in at-risk patients (–0.31 [95%CI: –0.57, –0.06] to –0.47 [95%CI: –0.82, –0.11]) and treating (–0.30 [95%CI:-0.57, –0.03] to –1.11 [95%CI: –1.47, –0.75]) AN, BN, and BED [51]. Further, binge reduction –0.66 [95%CI: –1.11, 0.22]) was also found in BED and BN patients [51]. While ICBT approaches were effective for up to 12 months, face-to-face CBT showed significant longer-term improvement (>12 months), especially in binge or purge frequency in BN (g=0.81, [95% CI 0.42 to 1.19], *p*<0.01) and BED (g=4.11, [95% CI 2.89 to 5.33], *p*<0.001) [49]. This suggests that while ICBT can be a valuable short-term treatment option, incorporating face-to-face therapy may enhance long-term outcomes and sustainability of recovery for individuals with EDs.

Another therapeutic modality commonly reported on in EDs is IDBI. IDBI was the alternative self-help intervention used in comparison to ICBT. Face-to-face IDBI demonstrates that these interventions are clinically significant, as shown by a meta-analysis containing 56 studies [52]. Dissonance-based prevention programs effectively reduce the thin ideal internalisation (*d* = 0.57), body dissatisfaction (*d* = 0.42), dieting (*d* = 0.37), negative affect (*d* = 0.29), and ED symptoms (*d* = 0.31) [52]. Similar results were found in this systematic review, with computer-based self-help IDBI being effective in reducing thin idealisation (*p<*0.01), body dissatisfaction (*p<*0.01), depression (*p<*0.01), and reward-based eating drive (*p*=0.045).

Whilst computer-based interventions appear effective with regards to the prevention of EDs, it was noted that there is a significant value in the therapeutic intervention, as evidenced by the greater effectiveness of guided versus non-guided self-help tools. Literature continues to demonstrate the importance of human interaction in symptom reduction, with included studies demonstrating greater effectiveness for guided self-help tools compared to non-guided [14, 30]. This is in line with the findings of this systematic review with peer– and therapist-led groups demonstrating higher participant satisfaction and more significant ED symptom reduction compared to the unguided computer-based self-help cognitive dissonance-based conditions [14]. Features identified in this systematic review, including engaging with peers, receiving guides from practitioners, and getting reminders improved patients’ experiences during treatment [53].

Studies included in this systematic review demonstrated equal outcome improvements for primary, secondary, and tertiary prevention compared to face-to-face interventions in all domains except for remission. One meta-analysis found that remission was present in 35.8% of face-to-face participants compared to 24.7% of participants in the computer-based group (RR=0.69, [95% CI 0.53 to 0.89], *p*=0.004, 4 RCTs, *n*=526) [54]. This indicates that computer-based self-help platforms, along with associated apps, can serve as effective early short-term preventative measures for patients awaiting face-to-face intervention. This approach not only reduces symptoms in all preventative tiers but also promotes ‘waiting well,’ providing valuable support for patients until they can meet with a clinician.

Linardon (2020) found that the majority of participants continue to prefer face-to-face treatment, which is highlighted in this systematic review by high drop-out rates in computer-based self-help platforms ± associated apps conditions. However, 50-70% of participants showed a willingness to use computer-based treatments for current ED symptoms, highlighting the importance of a complementary approach. Motivation and reassurance from peers and clinicians play a crucial role in improving outcomes and reducing dropout rates for those using computer-based treatment [11, 14, 30, 39, 46]. Factors such as current treatment experiences (b=1.18 (SE=0.26), OR=3.24, [95% CI 1.94 to 5.42]), attitude to internet interventions (b=1.97 (SE 0.20), OR=7.15, [95% CI 4.84 to 10.58]), and stigma (b=0.47 (SE 0.14), OR=1.59, [95% CI 1.22 to 2.08]) were demonstrated as significant contributing factors to the continuation of using the computer-based treatments [55].

### Comparison between unguided computer-based self-help platforms ± associated app modalities

This systematic review included various computer-based self-help platforms and associated apps that employ different approaches, all of which effectively target and reduce eating disorder symptoms and related behaviours [14, 31, 40, 46]. The studies reporting on outcomes indicated significant improvements. ICBT which focused on media internalisation was demonstrated more effective in reducing global ED pathology and improving quality of life compared to other approaches mentioned in this systematic review [39]. Aligned with this systematic review, the wider literature found that CBT-based short-term interventions improved the quality of life of participants with problematic social media use [56]. This can be explained in terms of the use of media focused on thinness culture and body dissatisfaction being demonstrated as a risk factor for ED development [57, 58]. Moreover, ICBT provides information which educates and aims to change maladaptive behaviours, which therefore targets ED symptoms and related behaviours [39]. With the increased social media use, media-embedded clinical practice and education are needed to ensure a limited impact on media-related risk factors such as the drive for thinness and body dissatisfaction [57]. While ICBT is most effective in managing maladaptive behaviours in all groups, IDBI is most effective in reducing eating pathology, especially in the global majority groups compared to white participants [31, 33].

IDBI is found to be most effective when managing specific behaviours, such as media internalisation; however, it does not target other psychopathologies, such as anorectic behaviours. Further, IDBI showed a higher effect size in thin-ideal internalisation compared to other outcomes, such as dieting, negative affect, and body dissatisfaction. These IDBI outcomes had smaller effect sizes than ICBT, suggesting that while IDBI may still provide some benefits, ICBT is generally more effective in achieving meaningful improvements in treatment outcomes.

This systematic review also found that computer-based interventions, such as psychoeducation, ICBT, and IDBI, also improved depression and negative affect. Face-to-face CBT that focuses on negative affect has been used to improve body shape and weight concerns in BN [59]. This approach helps individuals develop healthier coping strategies, enhance emotional regulation, and challenge distorted beliefs about body image, ultimately leading to more positive self-perceptions and reduced disordered eating behaviours. Depression and negative affect are associated with the development of maladaptive eating behaviours, which could develop into EDs [60]. Negative affect, alongside body dissatisfaction and thin-ideal internalisation, was also shown as a potential risk factor for disordered eating particularly in the Asian population, which could be explained by differences in body image culture [61].

The included samples included a significant number of participants if Asian descent, which could therefore explain the treatment effectiveness discrepancies [31, 33]. A research showed people of Asian descent have lower body satisfaction compared to those from European descent, a greater desire for thinness, a more significant concern with weight, and dissatisfaction with certain body parts [62]. In addition, women from Japan have a higher body dissatisfaction compared to any other East Asian countries, where Western media was one of the factors influencing their ideal body size [58]. It is therefore, possible that Asian participants experienced more psychological discomfort while being urged to question thin ideals, which resulted in superior intervention responses. Therefore, future studies and clinical practice should account for the impact of ethnicities on treatment effectiveness.

### Drop-out rates

Despite the effectiveness, the results must be cautiously interpreted, due to the risk of bias concerns found in the studies. Most included studies have a follow-up period of 12 months or less and have a high drop-out rate, with a mean drop-out rate of 39.4%, with one study not mentioning the drop-out rate [41], despite AG and YYK reaching out to the corresponding author to gather this data. The high dropout rates resulted in concerns regarding the risk of bias, which was reflected in the Cochrane RoB 2 judgment. Additionally, the high dropout rates may be attributed to the young participant sample sizes, as younger individuals tend to have a greater likelihood of disengaging. This can be explained by factors such as lack of motivation, time constraints, educational commitments, novelty seeking, previous treatment experiences, and a preference for different treatment formats [11, 32, 63]. Others argued drop-out rates were caused by unmet participants expectancy (OR=0.91, [95% CI 0.82 to 0.97]) and lower body mass index (OR=1.10, [95% CI 1.03 to 1.18]) [63]. These findings suggest that when participants’ anticipations regarding treatment outcomes are not fulfilled, or when they have a lower body mass index, they may be more likely to disengage from the program, highlighting the importance of managing expectations and addressing individual needs in treatment.

Two studies showed a drop-out rate of less than 20% [14, 45]. Stice explained retention methods including participant incentives, newsletters, and frequent follow-ups. Future researchers and clinicians should consider utilising short 5-minute telephone conversations to encourage continuing using the program which significantly improved participant adherence (T=-3.015, df =124, *p*=0.003) [64].

### Implications

This systematic review highlights the clinical effectiveness of unguided computer-based self-help platforms ± associated apps in the tiered healthcare system. Although the effectiveness varies, this study reports that the participants, the majority of whom were female, represented a diverse range of ethnicities, including individuals of American, European, Oceanian, and Asian descent. This diversity enhances the generalizability of the findings and underscores the need for culturally tailored interventions that address the unique experiences and challenges faced by different groups.

However, as most studies demonstrate effectiveness over short follow-up periods, additional research should be conducted to evaluate the effectiveness over a longer period with decreased drop-out rates. Furthermore, while unguided self-help platforms may demonstrate short-term effectiveness, they are not recommended as substitutes for in-person treatment. However, they can play a valuable role in specific situations, particularly as a primary intervention, and their scalability makes them a convenient option in many cases. Additionally, computer-based self-help platforms serve as a valuable supportive tool for individuals awaiting in-person treatments, in a ‘waiting well’ approach.

As the attrition rates were higher in the younger populations, it is recommended that additional research is conducted to evaluate manners that retain younger users.

### Future Considerations

Further studies need to be conducted to explore the effectiveness of the unguided computer-based self-help platforms ± associated apps in other populations, such as the male populations and across different ethnicities, in different ED types, such as in anorexia nervosa and avoidant/restrictive food intake disorder; discussing additional ED-related behaviours, such as perfectionism, and guilt. Additionally, it would be helpful to have longitudinal studies which examine effectiveness of these tools.

### Strengths

This systematic review has several strengths. Firstly, this systematic review found the effectiveness of different unguided computer-based self-help platforms ± associated apps as a potential temporary treatment for EDs. Secondly, the grouping of primary, secondary, and tertiary prevention clearly demonstrates its efficacy at different stages of prevention. Thirdly, the overall female-to-male ratio is representative of the prevalence of EDs worldwide. Lastly, this systematic review did not limit participant age, which improves the scope of interventions.

### Limitations

Despite the numerous strengths, this systematic review has some notable limitations. Firstly, most studies had significant dropout rates, which could have influenced the outcome interpretations. Secondly, although most studies had good sample sizes, future studies should aim to include greater sample sizes to improve reliability and validity of the findings. Lastly, some studies had missing data, and although personal communications with several authors had been made, no responses were given, except for the study by Stice (2020).

### Conclusion

Overall, unguided primary, secondary, and tertiary prevention computer-based self-help platforms ± associated apps are effective in reducing weight and body concerns, thin idealisation, binge eating, global ED pathology, and depression, which is sustained over a 12-month period. IDBI and ICBT were the most commonly used approaches and demonstrated the greatest effectiveness. This systematic review sustains that self-help platforms can be utilised to prevent the onset and the worsening of ED symptoms. However, the result needs to be interpreted cautiously, as the adherence to the intervention was low.

## Supporting information

Supportive statements

## Data Availability

The results of the datasets used in this systematic review are available from the corresponding author upon reasonable request.

https://doi.org/10.2196/preprints.60165

